# The AI Agent in the Room: Informing Objective Decision Making at the Transplant Selection Committee

**DOI:** 10.1101/2024.12.06.24318575

**Authors:** Bima J. Hasjim, Ghazal Azafar, Frank Lee, Tayyab S. Diwan, Shilpa Raju, Jed Adam Gross, Aman Sidhu, Hirohito Ichii, Rahul G. Krishnan, Muhammad Mamdani, Divya Sharma, Mamatha Bhat

## Abstract

**Importance:** Transplantation is one of the few areas in medicine where the definitive treatment is rationed. Subjective decision-making pose challenges towards the transplant selection process. It has been proposed that large language models (LLMs) as artificial intelligent (AI) agents could provide objectivity in decision-making to solve complex problems.

**Objective:** To examine the performance of a multidisciplinary selection committee of AI agents (AI-SC) as a proof-of-concept towards objectivity in the liver transplant (LT) selection process.

**Design:** The AI-SC consisted of four LLMs: transplant hepatologist, transplant surgeon, cardiologist, and social worker. Zero-shot prompting with chain-of thought was used. Decisions were made based on clinicodemographic characteristics at time of waitlisting and LT.

**Setting:** National LT cohort.

**Participants:** Adult patients receiving deceased donor LT from 2004-2023 were extracted from the Scientific Registry of Transplant Recipients (SRTR) and clinical vignettes were generated. Standard absolute contraindications to LT were randomly assigned to a subset of patients to expose the AI-SC to cases of patients declined for LT.

**Exposures:** Clinicodemographic characteristics at waitlisting and transplantation.

**Main Outcomes and Measures:** The AI-SC’s accuracy with either: 1) listing candidates if LT would offer a 6-month or 1-year survival benefit or 2) declining candidates if contraindications to LT are present or if LT would not offer those survival benefits.

**Results:** Of 8,412 patients, 83.6% were waitlisted and 16.4% had contraindications to LT. The AI-SC was able to accurately identify contraindications to LT (accuracy: 98.2%, 95%CI 97.9%-98.4%), predict 6-month (94.9%, 95%CI 94.4%-95.3%) and 1-year (92.0%, 95%CI 91.4%-92.6%) survival. HCC burden beyond Milan criteria was the most common reason for accepted patients who were declined by AI-SC (False Negative). Malignancy was the most common cause of death prior to 6-month or 1-year end points (False Positive). The AI-SC most frequently did not perceive a lack of social support or severe cardiopulmonary disease as barriers to LT.

**Conclusions and Relevance:** LLMs can be leveraged to simulate the LT-SC meetings and provide accurate, objective insights on patients who may or may not benefit from LT. Lessons learned from this proof-of-concept are a provocative step towards making the LT selection process more equitable and objective.

**Key Points:** *Question:* Can a multidisciplinary selection committee of artificial intelligence-based agents (AI-SC) accurately select liver transplant (LT) candidates based on potential survival benefit and contraindications to LT?

*Findings:* Clinical vignettes were generated from 8,412 LT candidates from the Scientific Registry of Transplant Recipients (SRTR). Of these, 16.4% were randomly assigned standard absolute contraindications to LT. The AI-SC (GPT-4, OpenAI) reviewed and selected LT candidates with accuracies of 98.2% in identifying contraindications to LT, 94.9% in predicting 6-month survival benefit, and 92.0% in predicting 1-year survival benefit.

*Meaning:* Multi-agent models may be leveraged to provide guidance towards objective decision-making in transplant candidacy.

## Introduction

Solid organ transplantation is one of the few areas in medicine where the definitive treatment is explicitly rationed because the demand far exceeds the available allograft supply. In the context of liver transplantation (LT), it is estimated that only 5-10% of all patients with end stage liver disease (ESLD) are placed on the waitlist and less than 5% of those were transplanted.^1–3^ The decision to place a patient on the waitlist is made by a multidisciplinary LT selection committee (LT-SC) based on the projected survival gain an allograft would offer.^4^ National organizations such as United Network for Organ Sharing and American Association for the Study of Liver Disease have established guidelines on its proceedings to ensure fairness, promote equitable access, and standardize selection practices across centers.^5–8^

Although these efforts have created some uniformity, decisions made by the LT-SC may still be prone to subjectivity and bias. For example, delineating “relative” and “absolute" contraindications may vary significantly by provider type, availability of resources, and center volume.^9^ Unwritten institutional policies, external pressures^10^, and ambiguity between advocacy and stewardship roles are a few barriers to effective group decision making.^7,11^ Further, implicit biases against particular psychosocial profiles may influence the LT-SC’s beliefs regarding a candidate’s ability to achieve a successful outcome.^12^ Consistency and fairness of LT-SC decisions may also be influenced by which specialists are represented and their familiarity with the patient. Reportedly, transplant providers tend to advocate more strongly for patients with whom they have long-standing relationships with.^7^ Inversely, discussions tended to be shorter and more negative in tone when there was no one to advocate for the patient.^7^ Lastly, the decision to determine who ultimately deserves a LT, and subsequently who lived and died, is inherently uncomfortable and may lead to emotional conclusions – even among experienced members of the team.^7^ Currently, the LT selection process remain largely unexplored as it is often kept behind closed doors to ensure patient privacy, but also to promote honest discussions that are complex, difficult, and high stakes.^7,12^

Large language models (LLMs) have emerged as a powerful tool within the medical field with great potential. Early applications of LLMs have been shown to excel in medical question-and-answer tasks^13,14^, perioperative surgical risk stratification^15^, and summarizing electronic health record data.^16,17^ LLMs may act as an autonomous artificial intelligent (AI) ‘agent’ to perform specialized tasks, but also engage in multi-agent collaborations, share information, and make decisions collectively to solve complex problems by simulating human interactions.^18–20^ In the business industry, AI agents are increasingly used to improve efficiency, automate processes, and effect quality improvement initiatives^21–23^, but its applications in the medical field are still limited.

Given its potential, multi-agent LLMs could be a powerful resource for evaluating LT candidates. We designed a selection committee of artificial intelligence-based agents (AI-SC) to mimic the multidisciplinary LT-SC and evaluated their performance in identifying candidates who were likely to achieve a survival benefit from LT. We also investigate the decision making processes and potential biases in the AI-SC. Such findings can offer insights into how AI may serve as a potential clinical decision support tool and provide guidance towards objective decision making in the LT-SC.

## Methods

### Dataset

This was a cohort study of the Scientific Registry of Transplant Recipients (SRTR) who received a deceased donor LT (DDLT) from 1/1/2004-6/1/2024. The SRTR is a national registry of all donor, waitlisted candidates, and transplant recipients submitted by members of the United States (US) Organ Procurement and Transplantation Network (OPTN).^24^ The Health Resources and Services Administration, US Department of Health and Human Services oversee activities of OPTN and SRTR contractors. This study was approved by the Institutional Review Board at University of California, Irvine (#4474), followed the Transparent Reporting of a Multivariable Prediction Model for Individual Prognosis or Diagnosis (TRIPOD) guidelines, and adhered strictly to OpenAI’s privacy policy.^25^

### Cohort Selection

Adult (≥18-years-old) DDLT patients with adequate post-LT follow-up were included in the study. Patients with history of a previous transplant, living donor LT, and pediatric patients were excluded as their candidacy and organ allocation processes differ from adult DDLT (*Figure S1*).

Since the SRTR does not report on rejected patients from the waitlist, a subset of patients from the SRTR (N=1,379, 16.4%) were randomly assigned absolute contraindications to LT (e.g., metastatic hepatocellular carcinoma (HCC), active extrahepatic malignancy, cardiopulmonary disease) as published by the literature.^26^ The distribution of active extrahepatic malignancy were based on the World Health Organization’s most prevalent types of cancer-related mortality: breast, colorectal, hematologic, lung, pancreatic, prostate, and gastric cancer.^27^

### Experimental Design: AI-agents in the AI-SC

The primary LLM was OpenAI’s gpt-4o-2024-08-06. CrewAI (v0.63.6), AgentOps (v0.3.12), and langchain_openai libraries in Python were used to develop the AI-SC.

*Figure 1* describes the overview of the experimental workflow. An AI medical scribe generated and presented clinical vignettes of all patients included in the analysis to the AI-SC, which consisted of four AI agents with specialized roles: transplant hepatologist, transplant surgeon, cardiologist, and social worker. The AI-SC was tasked with classifying each patient as a valid transplant candidate if they 1) had no contraindication to LT, 2) were predicted to benefit from LT with ≥6-month survival or 3) ≥1-year survival. Prompts were developed and refined to ensure domain-specific alignment and to engineer purpose-built prompts specifically optimized for the AI-SC (*Figure S2*). Alternative prompting strategies and their performance were conducted on 250 randomly-selected patients (*Table S1*).

**Figure 1.**
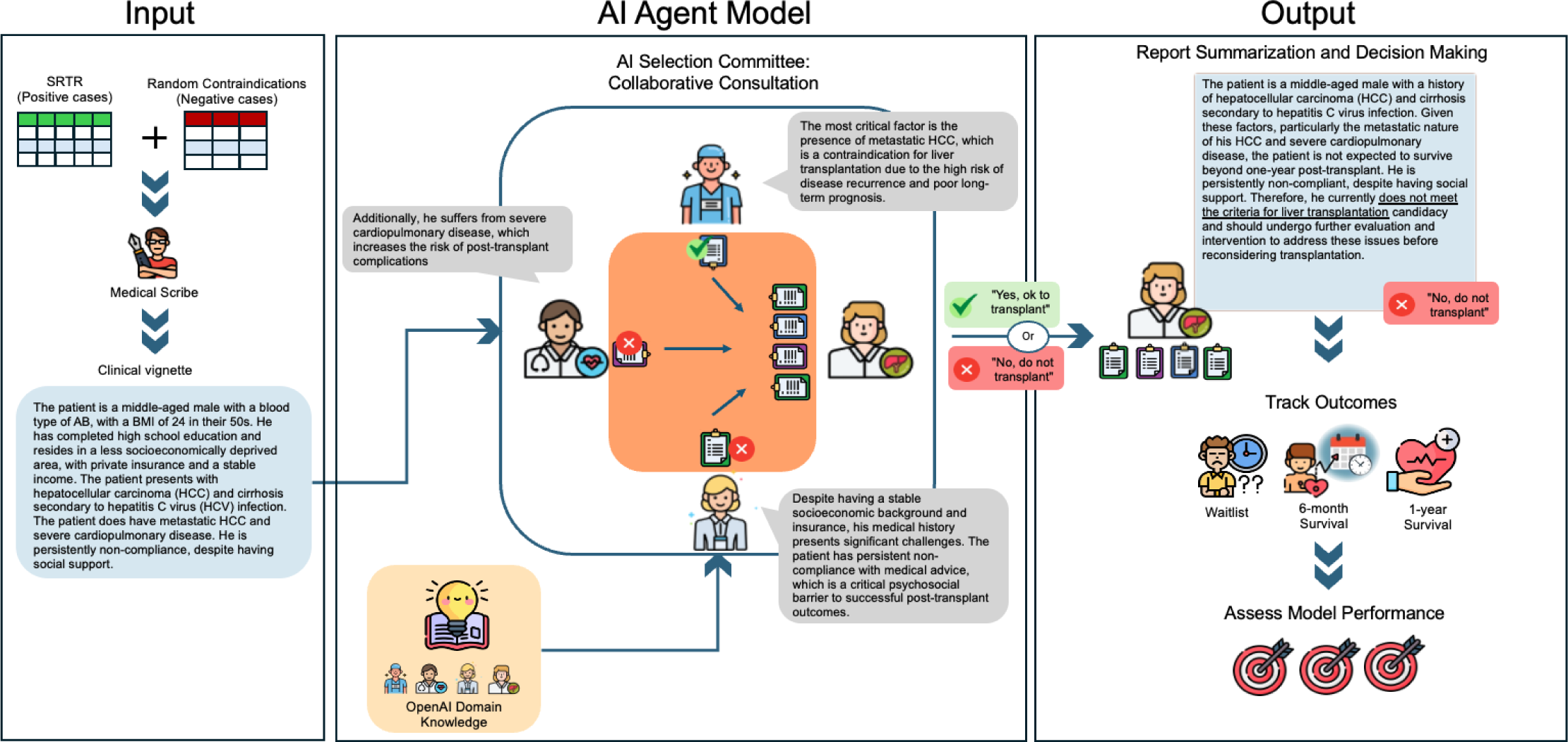
Overview of experimental workflow. Clinical vignettes, derived from SRTR tabular data containing patient characteristics, clinical history, and laboratory results, were provided to a LLM configured as a medical writer. The generated vignettes were subsequently reviewed by a multi-disciplinary AI agent committee that were tasked with evaluating the patient’s transplant eligibility based on the presence of absolute contraindications to LT or the likelihood of 6-month, or 1-year survival benefit. Each agent was prompted to function within a defined role, break up the task by each agent’s expertise with domain knowledge inherent to OpenAI, and to think step-by-step (zero-shot with chain of thought prompting (zero-shot-COT).^58^ Consensus on listing recommendations was reached through a structured process of report sharing and analysis with the AI transplant hepatologist. The AI transplant hepatologist, responsible for synthesizing the reports and speaking on behalf of the AI-SC, was instructed to provide a binary “Yes, ok to transplant” or “No, do not transplant” answer with a detailed report explaining the basis of the decision. The performance of the AI-SC decisions were compared to reported outcomes in SRTR. Figure 1 illustrates the experimental workflow for a fictional patient profile.

### Outcomes and Clinical Variables

The primary objective of this study was to assess the accuracy of AI-SC in predicting 1-year post-LT survival based on clinicodemographic characteristics reported at waitlisting and transplantation. Secondary objectives include identifying contraindications to LT and predicting 6-month post-LT survival.

A total of 59, equally-weighted, variables were considered and included clinicodemographic, social determinants of health (SDOH), and ESLD characteristics. The Area Deprivation Index (ADI) was merged with the SRTR as a measure of SDOH and identified patients residing in disadvantaged ZIP codes in the US (*Supplemental Methods*).^28,29^ Redundant variables (e.g., history of diabetes vs. insulin dependence) and those with >20% missing data were excluded^30,31^ to minimize hallucinations. A temperature setting of 0.1 was also used to mitigate hallucinations.^32,33^ A random sample of 500 vignettes were manually reviewed (BJH and FGL) and hallucinations were identified in <1% of cases.

### Cosine Similarity Index and Fairness

Cosine similarity index analysis identified the prevalent variables used by each individual member of the AI-SC for their decisions and the top 20 variables with the highest cosine similarity index were reported.^34–37^ Disparate Impact (DI) analysis assesses the fairness of AI-SC decisions by quantifying the likelihood of a positive classification (“Yes, ok to transplant”) for a subgroup relative to others.^38^ DI>1 denotes the subgroup is advantaged by the AI-SC as it has a higher proportion of the positive classification compared to other groups. When DI<1, the subgroup of interest is disadvantaged by the AI-SC since the subgroup had a lower proportion of the positive classification compared to other groups.^38^ A threshold of DI≥1.2 or DI≤0.8 signifies unfair classifications.^38^ Additional details on these analyses and calculations can be found in the *Supplemental Methods*.

### Statistical Analyses and Performance Evaluation

AI-SC performance was evaluated by accuracy, specificity, recall, precision, and F1-score. 95% Confidence Intervals (95%CI) were estimated using 2,500 bootstrap iterations.^15^ False Positive (FP) cases were defined as those classified as “Yes, ok to transplant” (likely benefit past 6-months or 1-year) but died before the clinical endpoint (assessed by cause of death). For cohorts with absolute contraindications to LT, FP cases were those who were classified as “Yes, ok to transplant” despite the presence of an absolute contraindication to LT. False Negative (FN) cases were defined as those who were transplanted or survived past the clinical endpoint, but were rejected from LT (“No, do not transplant”) by the AI-SC due to the perceived presence of contraindications or the belief that they would not survive 6-months or 1-year after LT (assessed by manual review of AI-SC reports).

Descriptive analyses and summary statistics of clinicodemographic covariates were reported with means and standard deviation (SD) or medians and interquartile ranges (IQR). Kruskal-Wallis rank sum test and the Pearson’s chi-square test for independence were used to compare continuous and categorical variables, respectively. Two-sided p-values were used and an alpha level <0.05 were considered statistically significant. Data processing and analysis were performed using R studio (version 4.2.3, Boston, MA, US) and Python (version 3.10.12, Beaverton, OR, US).

## Results

There were 8,412 patients included in the analysis: 83.6% (N=7,033) were waitlisted and 16.4% (N=1,379) had contraindications to LT. The median (IQR) age was 58-years-old (53.0, 62.0 years) with a median follow up of 8.5 years. In the overall cohort, 21.9% were female, 66.1% were non-Hispanic White, 9.5% were non-Hispanic Black, 15.3% were Hispanic, 8.0% were Asian, and 1.1% were Other. The majority of patients had private insurance (59.1%), independent functional status (74.9%), and most often resided in moderately SES deprived areas (25.8%). The 6-month and 1-year mortality rate was 4.0% and 7.5%, respectively (*Table 1*).

**Table 1.**
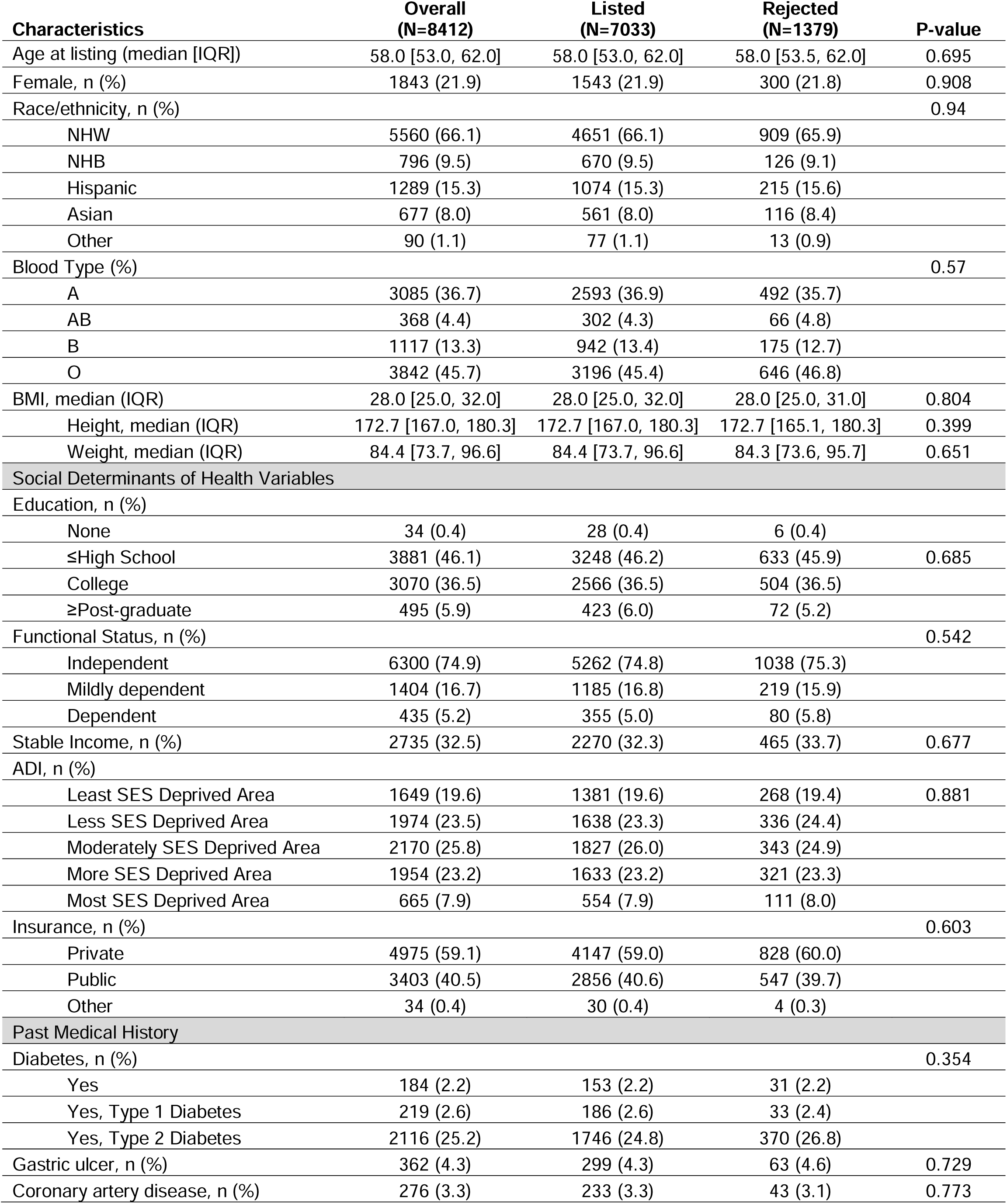
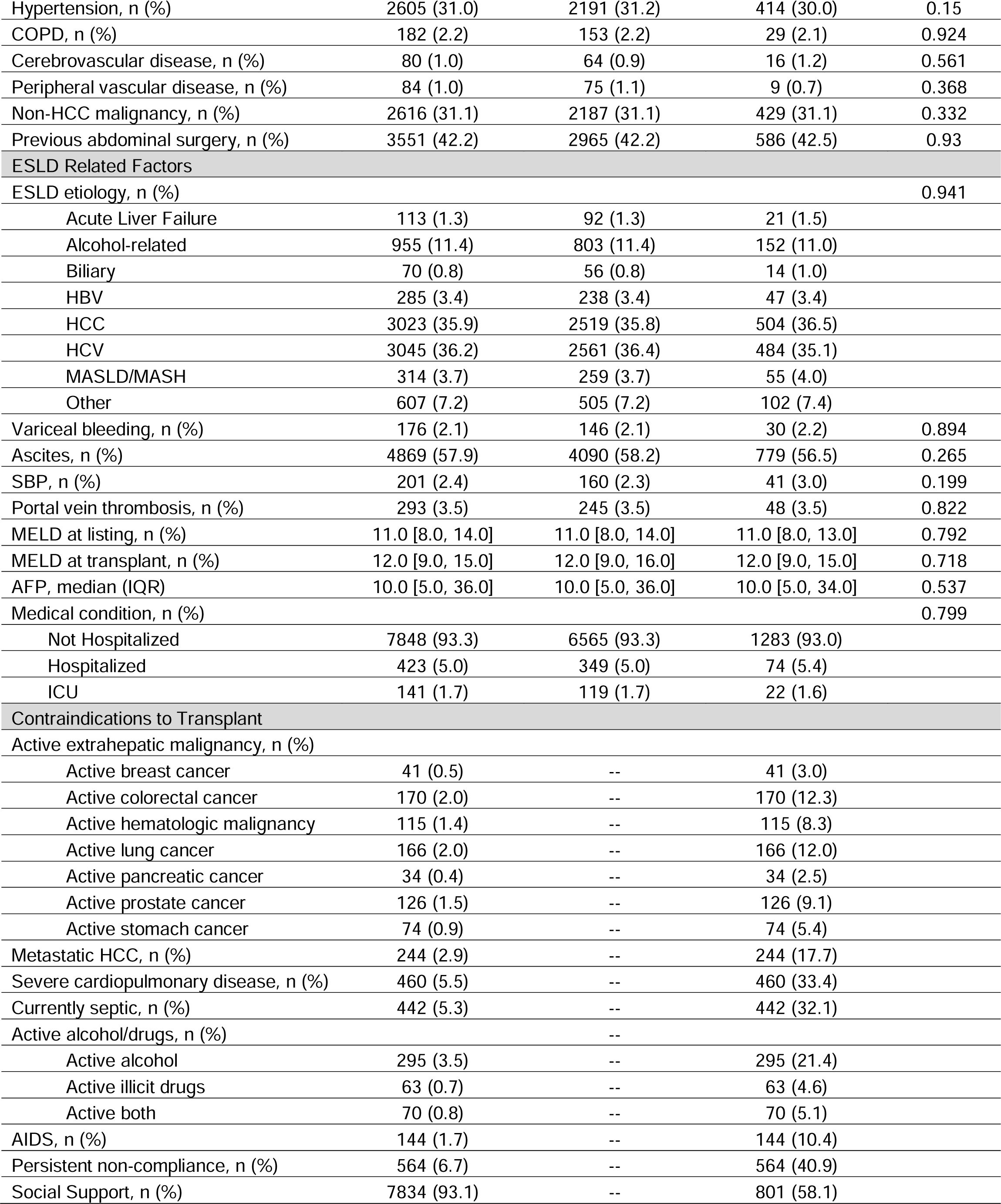
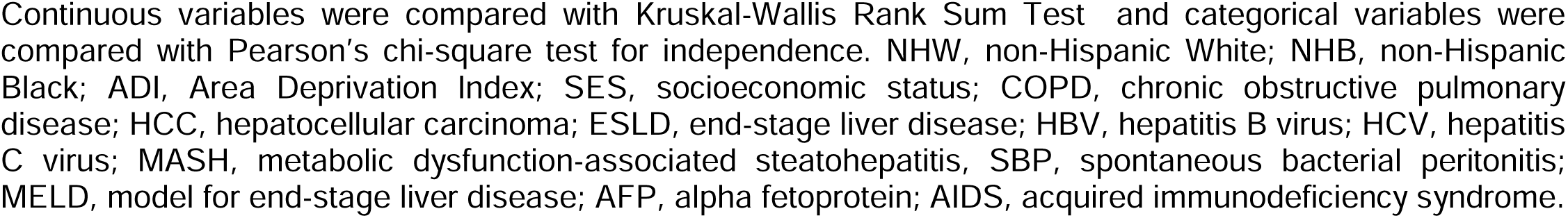
Baseline demographics and clinical characteristics of liver transplant recipients in the study.

| Characteristics | Overall<br>(N=8412) | Listed<br>(N=7033) | Rejected<br>(N=1379) | P-value |
| --- | --- | --- | --- | --- |
| Age at listing (median [IQR]) | 58.0 [53.0, 62.0] | 58.0 [53.0, 62.0] | 58.0 [53.5, 62.0] | 0.695 |
| Female, n (%) | 1843 (21.9) | 1543 (21.9) | 300 (21.8) | 0.908 |
| Race/ethnicity, n (%) |  |  |  | 0.94 |
| NHW | 5560 (66.1) | 4651 (66.1) | 909 (65.9) |  |
| NHB | 796 (9.5) | 670 (9.5) | 126 (9.1) |  |
| Hispanic | 1289 (15.3) | 1074 (15.3) | 215 (15.6) |  |
| Asian | 677 (8.0) | 561 (8.0) | 116 (8.4) |  |
| Other | 90 (1.1) | 77 (1.1) | 13 (0.9) |  |
| Blood Type (%) |  |  |  | 0.57 |
| A | 3085 (36.7) | 2593 (36.9) | 492 (35.7) |  |
| AB | 368 (4.4) | 302 (4.3) | 66 (4.8) |  |
| B | 1117 (13.3) | 942 (13.4) | 175 (12.7) |  |
| O | 3842 (45.7) | 3196 (45.4) | 646 (46.8) |  |
| BMI, median (IQR) | 28.0 [25.0, 32.0] | 28.0 [25.0, 32.0] | 28.0 [25.0, 31.0] | 0.804 |
| Height, median (IQR) | 172.7 [167.0, 180.3] | 172.7 [167.0, 180.3] | 172.7 [165.1, 180.3] | 0.399 |
| Weight, median (IQR) | 84.4 [73.7, 96.6] | 84.4 [73.7, 96.6] | 84.3 [73.6, 95.7] | 0.651 |
| Social Determinants of Health Variables |  |  |  |  |
| Education, n (%) |  |  |  |  |
| None | 34 (0.4) | 28 (0.4) | 6 (0.4) |  |
| ≤High School | 3881 (46.1) | 3248 (46.2) | 633 (45.9) | 0.685 |
| College | 3070 (36.5) | 2566 (36.5) | 504 (36.5) |  |
| ≥Post-graduate | 495 (5.9) | 423 (6.0) | 72 (5.2) |  |
| Functional Status, n (%) |  |  |  | 0.542 |
| Independent | 6300 (74.9) | 5262 (74.8) | 1038 (75.3) |  |
| Mildly dependent | 1404 (16.7) | 1185 (16.8) | 219 (15.9) |  |
| Dependent | 435 (5.2) | 355 (5.0) | 80 (5.8) |  |
| Stable Income, n (%) | 2735 (32.5) | 2270 (32.3) | 465 (33.7) | 0.677 |
| ADI, n (%) |  |  |  |  |
| Least SES Deprived Area | 1649 (19.6) | 1381 (19.6) | 268 (19.4) | 0.881 |
| Less SES Deprived Area | 1974 (23.5) | 1638 (23.3) | 336 (24.4) |  |
| Moderately SES Deprived Area | 2170 (25.8) | 1827 (26.0) | 343 (24.9) |  |
| More SES Deprived Area | 1954 (23.2) | 1633 (23.2) | 321 (23.3) |  |
| Most SES Deprived Area | 665 (7.9) | 554 (7.9) | 111 (8.0) |  |
| Insurance, n (%) |  |  |  | 0.603 |
| Private | 4975 (59.1) | 4147 (59.0) | 828 (60.0) |  |
| Public | 3403 (40.5) | 2856 (40.6) | 547 (39.7) |  |
| Other | 34 (0.4) | 30 (0.4) | 4 (0.3) |  |
| Past Medical History |  |  |  |  |
| Diabetes, n (%) |  |  |  | 0.354 |
| Yes | 184 (2.2) | 153 (2.2) | 31 (2.2) |  |
| Yes, Type 1 Diabetes | 219 (2.6) | 186 (2.6) | 33 (2.4) |  |
| Yes, Type 2 Diabetes | 2116 (25.2) | 1746 (24.8) | 370 (26.8) |  |
| Gastric ulcer, n (%) | 362 (4.3) | 299 (4.3) | 63 (4.6) | 0.729 |
| Coronary artery disease, n (%) | 276 (3.3) | 233 (3.3) | 43 (3.1) | 0.773 |
| Hypertension, n (%) | 2605 (31.0) | 2191 (31.2) | 414 (30.0) | 0.15 |
| COPD, n (%) | 182 (2.2) | 153 (2.2) | 29 (2.1) | 0.924 |
| Cerebrovascular disease, n (%) | 80 (1.0) | 64 (0.9) | 16 (1.2) | 0.561 |
| Peripheral vascular disease, n (%) | 84 (1.0) | 75 (1.1) | 9 (0.7) | 0.368 |
| Non-HCC malignancy, n (%) | 2616 (31.1) | 2187 (31.1) | 429 (31.1) | 0.332 |
| Previous abdominal surgery, n (%) | 3551 (42.2) | 2965 (42.2) | 586 (42.5) | 0.93 |
| **ESLD Related Factors** |  |  |  |  |
| ESLD etiology, n (%) |  |  |  | 0.941 |
| Acute Liver Failure | 113 (1.3) | 92 (1.3) | 21 (1.5) |  |
| Alcohol-related | 955 (11.4) | 803 (11.4) | 152 (11.0) |  |
| Biliary | 70 (0.8) | 56 (0.8) | 14 (1.0) |  |
| HBV | 285 (3.4) | 238 (3.4) | 47 (3.4) |  |
| HCC | 3023 (35.9) | 2519 (35.8) | 504 (36.5) |  |
| HCV | 3045 (36.2) | 2561 (36.4) | 484 (35.1) |  |
| MASLD/MASH | 314 (3.7) | 259 (3.7) | 55 (4.0) |  |
| Other | 607 (7.2) | 505 (7.2) | 102 (7.4) |  |
| Variceal bleeding, n (%) | 176 (2.1) | 146 (2.1) | 30 (2.2) | 0.894 |
| Ascites, n (%) | 4869 (57.9) | 4090 (58.2) | 779 (56.5) | 0.265 |
| SBP, n (%) | 201 (2.4) | 160 (2.3) | 41 (3.0) | 0.199 |
| Portal vein thrombosis, n (%) | 293 (3.5) | 245 (3.5) | 48 (3.5) | 0.822 |
| MELD at listing, n (%) | 11.0 [8.0, 14.0] | 11.0 [8.0, 14.0] | 11.0 [8.0, 13.0] | 0.792 |
| MELD at transplant, n (%) | 12.0 [9.0, 15.0] | 12.0 [9.0, 16.0] | 12.0 [9.0, 15.0] | 0.718 |
| AFP, median (IQR) | 10.0 [5.0, 36.0] | 10.0 [5.0, 36.0] | 10.0 [5.0, 34.0] | 0.537 |
| Medical condition, n (%) |  |  |  | 0.799 |
| Not Hospitalized | 7848 (93.3) | 6565 (93.3) | 1283 (93.0) |  |
| Hospitalized | 423 (5.0) | 349 (5.0) | 74 (5.4) |  |
| ICU | 141 (1.7) | 119 (1.7) | 22 (1.6) |  |
| **Contraindications to Transplant** |  |  |  |  |
| Active extrahepatic malignancy, n (%) |  |  |  |  |
| Active breast cancer | 41 (0.5) | -- | 41 (3.0) |  |
| Active colorectal cancer | 170 (2.0) | -- | 170 (12.3) |  |
| Active hematologic malignancy | 115 (1.4) | -- | 115 (8.3) |  |
| Active lung cancer | 166 (2.0) | -- | 166 (12.0) |  |
| Active pancreatic cancer | 34 (0.4) | -- | 34 (2.5) |  |
| Active prostate cancer | 126 (1.5) | -- | 126 (9.1) |  |
| Active stomach cancer | 74 (0.9) | -- | 74 (5.4) |  |
| Metastatic HCC, n (%) | 244 (2.9) | -- | 244 (17.7) |  |
| Severe cardiopulmonary disease, n (%) | 460 (5.5) | -- | 460 (33.4) |  |
| Currently septic, n (%) | 442 (5.3) | -- | 442 (32.1) |  |
| Active alcohol/drugs, n (%) |  | -- |  |  |
| Active alcohol | 295 (3.5) | -- | 295 (21.4) |  |
| Active illicit drugs | 63 (0.7) | -- | 63 (4.6) |  |
| Active both | 70 (0.8) | -- | 70 (5.1) |  |
| AIDS, n (%) | 144 (1.7) | -- | 144 (10.4) |  |
| Persistent non-compliance, n (%) | 564 (6.7) | -- | 564 (40.9) |  |
| Social Support, n (%) | 7834 (93.1) | -- | 801 (58.1) |  |

Continuous variables were compared with Kruskal-Wallis Rank Sum Test and categorical variables were compared with Pearson's chi-square test for independence. NHW, non-Hispanic White; NHB, non-Hispanic Black; ADI, Area Deprivation Index; SES, socioeconomic status; COPD, chronic obstructive pulmonary disease; HCC, hepatocellular carcinoma; ESLD, end-stage liver disease; HBV, hepatitis B virus; HCV, hepatitis C virus; MASH, metabolic dysfunction-associated steatohepatitis, SBP, spontaneous bacterial peritonitis; MELD, model for end-stage liver disease; AFP, alpha fetoprotein; AIDS, acquired immunodeficiency syndrome.

### AI-SC Performance

The performance of the AI-SC is presented in Figure 2A. The accuracy of identifying absolute contraindications to LT was 98.2% (95%CI 97.9%-98.4%). When determining clinical benefit of 6-month and 1-year survival, the accuracy was 94.9% (95%CI 94.4%-95.3%) and 92.0% (95%CI 91.4%-92.6), respectively. Sample cases are listed in *Table S2*. Among FP cases, the most frequent cause of death were infections at 6-months (21.7%) and malignancy at 1-year (28.5%) (Figure 2B). When reviewing cases with absolute contraindications to LT, the agents most frequently did not perceive a lack of social support (32.3%) or severe cardiopulmonary disease (33.1%) as "absolute" barriers to transplantation (Figure 2C). The most common reason for rejection in FN cases were tumors outside of the Milan Criteria (53.8% contraindications to LT; 53.8% 6-month survival; 60.9% 1-year survival) (Figure 2D).

**Figure 2.**
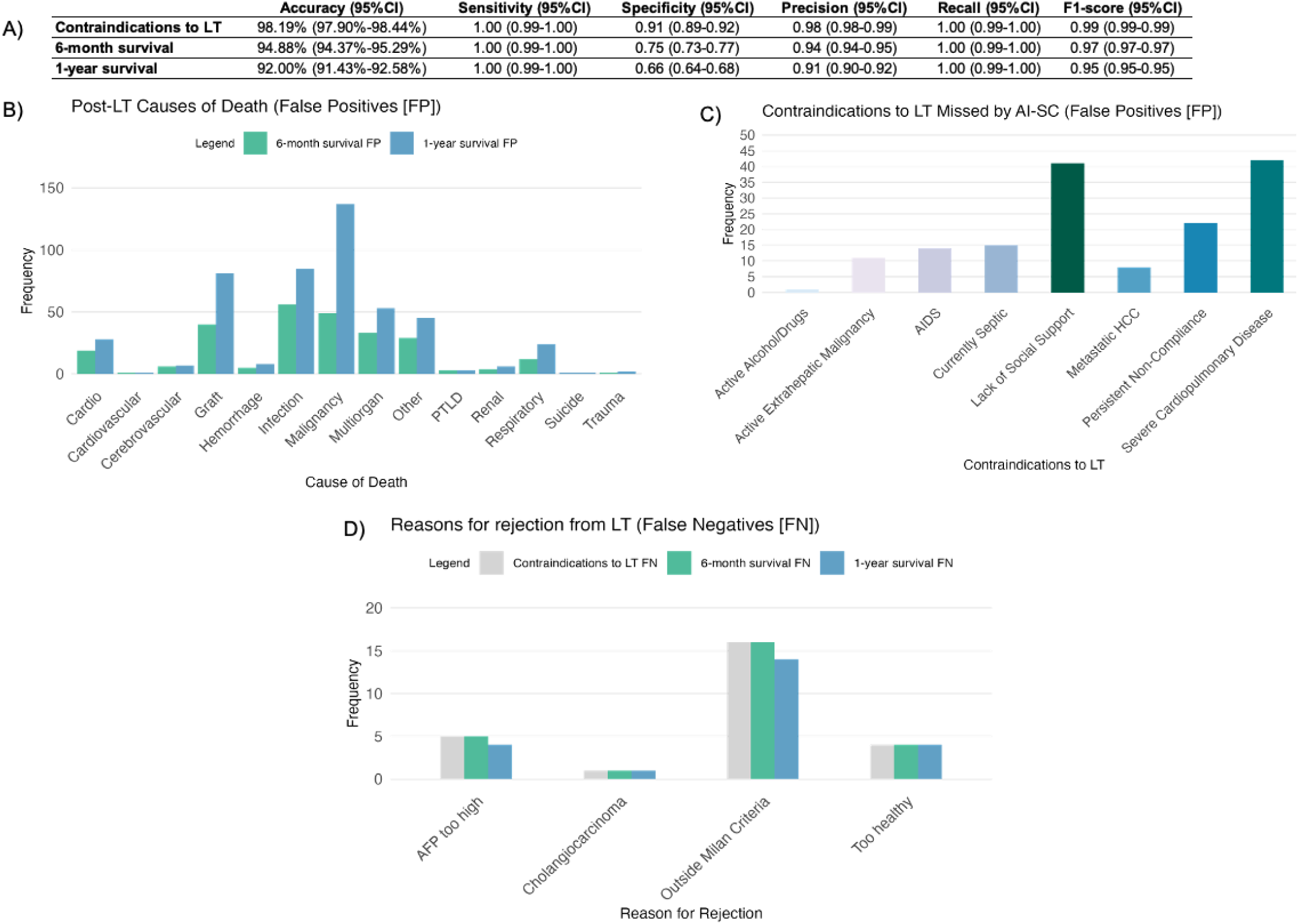
Performance of the model across different clinical endpoints. A) Performance of the model across different clinical endpoints. B) When assessing False Positives (patients who were deemed to likely benefit past 6-months or 1-year but died before the clinical endpoint), the most frequent cause of death was malignancy. C) Among cases with absolute contraindications to LT, the AI-SC most frequently didn’t perceive a lack of social support or severe cardiopulmonary disease as barriers to transplantation. D) When assessing False Negatives (patients that were rejected by the AI-SC due to contraindications or the belief that they wouldn’t survive past 6-months or 1-year after transplant but were transplanted and lived past that end point) the most common reason for rejection were tumors outside of the Milan Criteria.

### Cosine Similarity Index and Fairness Analyses

ESLD primary diagnosis and recipient age were among the top 5 most important variables used by all members of the AI-SC. Each AI specialist used different variables in their decision making and report generation based on their roles. Notable covariates used by the AI social worker included: persistent non-compliance, insurance type, ADI quintile category, and education. The AI cardiologist focused on variables such as history of cardiovascular disease, hypertension, coronary artery disease, and pulmonary embolism. Unique covariates used by the AI transplant surgeon were HCC tumor size, extrahepatic malignancy and spread. The AI transplant hepatologist had the most overlap between other specialists, but unique variables noted were sex and status on life support (Figure 3). The AI-SC were collectively fair with slight disadvantages (0.8<DI<1) observed for female, multiracial, Hispanic, less SES deprived areas, grade school education, and biliary ESLD etiology (Figure 4).

**Figure 3.**
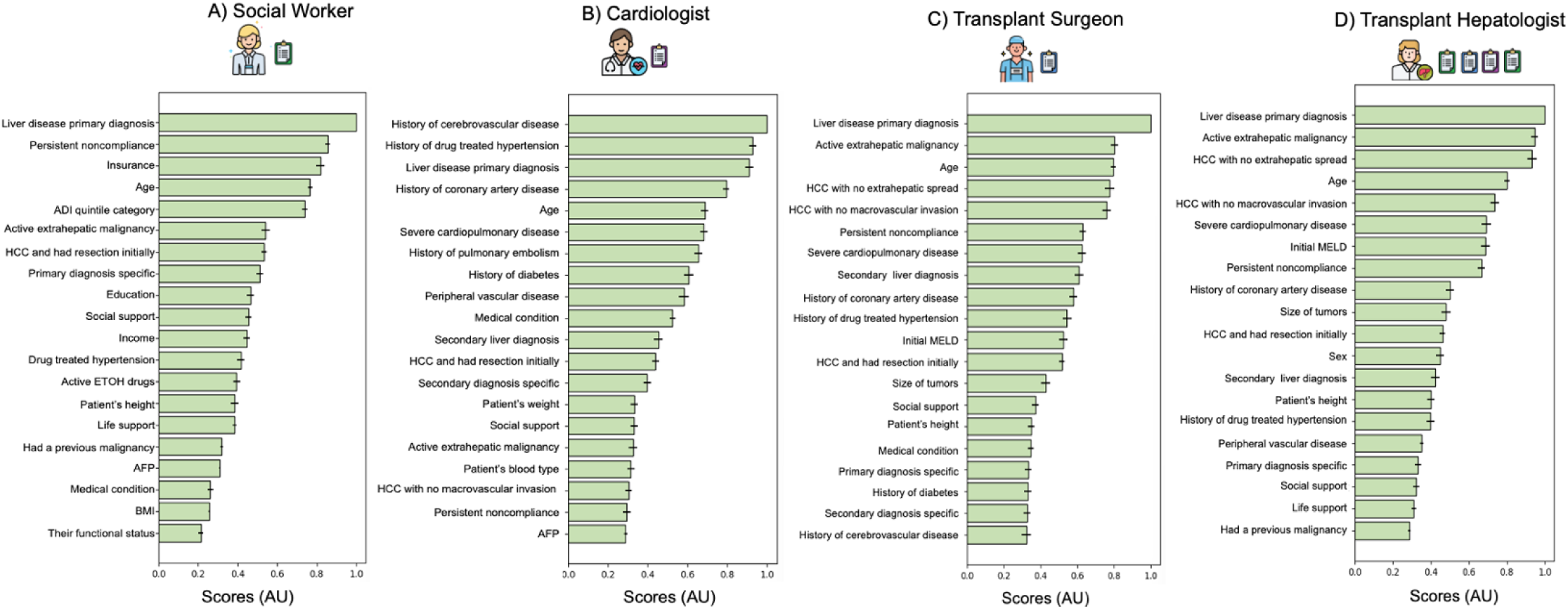
Cosine similarity index analysis of reports from each AI-SC committee members. This scoring system enables for an objective assessment of whether each AI agent was appr addressing relevant variables. Cosine similarity index analyses show AI clinical-specialty nuance priately in the variables used for decision making for the A) social worker, B) cardiologist, C) transplant surgeon, and D) transplant hepatologist.

**Figure 4.**
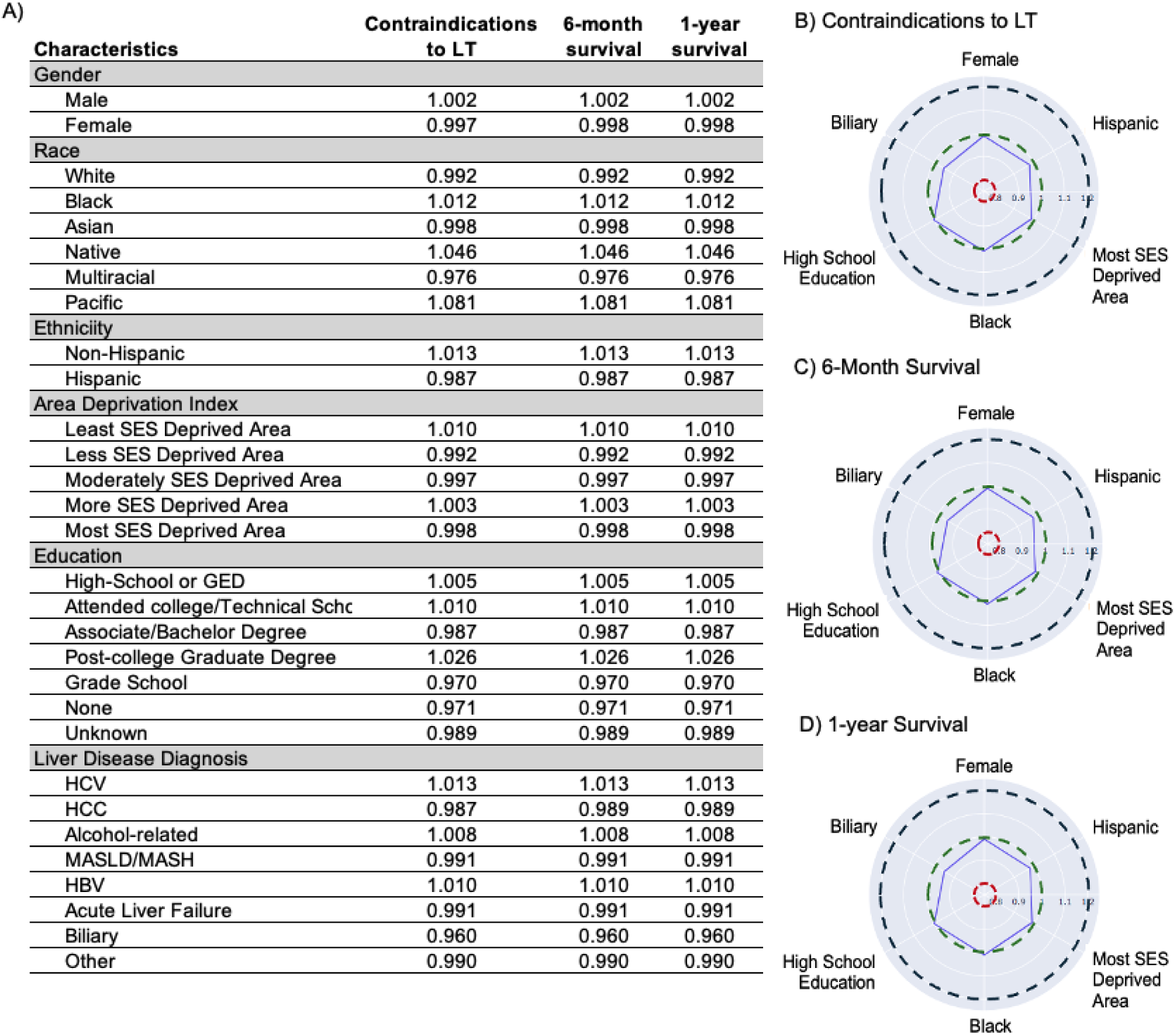
Disparate impact analyses for fairness of AI-SC when determining patients deemed worthy for transplantation when assessing contraindications to transplant, projected 6-month and 1-year survival. A) DI analyses for fairness of AI-SC when determining patients deemed worthy for transplantation when assessing contraindications to transplant, projected 6-month and 1-year survival. When DI>1, the subgroup of interest is advantaged as it has a higher proportion of positive outcomes compared to the other groups. When DI<1, the subgroup of interest is disadvantaged since the subgroup had a lower proportion of positive outcomes compare to the other groups. Spider plots of DI for historically-disadvantaged subgroups when B) identifying contraindications to LT, C) predicting 6-month survival, and D) 1-year survival.

## Discussion

Despite established guidelines, there is wide variability in LT selection practice patterns among and within LT centers.^5–8^ In the present study, LLMs can be leveraged to simulate the LT-SC meetings through AI agents and provide accurate, objective insights towards LT candidates. The AI-SC were able to accurately identify contraindications to LT (98.2%) and predict 6-month (94.8%) and 1-year (92.0%) post-LT survival. Lessons learned from our study are a provocative step towards objective decision making for transplant candidates and standardization of the selection process.

The rapid advancements in LLM technology and agentic AI have generated excitement over its potential clinical applications.^39^ In our study, agentic reasoning capabilities of LLMs can simulate LT-SC discussions and projected survival benefit. Similarly, Ferber et al. developed a clinical, decision-support AI tool for gastrointestinal cancer cases using AI agents.^40^ These agents can draw the correct conclusions in 93.6% of cases utilizing multimodal information (e.g., text, radiology, document retrieval) and provide helpful recommendations grounded by relevant medical literature in 82.5% of cases.^40^ In another study, the use of an AI agent companion improved clinical decision-making quality in 63.7% of real-world complex cases and those who used the AI agents performed superiorly to those who did not.^41^ AI agents may mitigate sources of inconsistency in the transplant selection process by providing an objective voice grounded in patient data and making subspecialty knowledge accessible when unavailable.^41^ The present study may serve as a proof-of-concept for more advanced LLM-agents in clinical decision support tools.

There are multiple scenarios in which AI integration in the LT selection process could be pursued. In one example, the AI-SC may provide a preliminary decision that is reviewed by the LT-SC. Alternatively, a LT-SC can decide upfront, followed by solicitation from the AI model. In all scenarios, human vigilance is paramount as hallucinations and bias are inherent dangers in LLMs and the burden of liability rests on the LT-SC.^42,43^ Although the AI-SCs were able to make fair assessments on transplant candidacy in our study, current LLM models are still prone to bias against historically-disadvantaged populations and others. Zack et al., found that GPT-4’s medical decision-making were significantly associated with demographic attributes and were less likely to recommend advanced imaging and intervention to Black and female patients, respectively.^44^ LLMs and other machine learning (ML) models can propagate and amplify harmful societal biases.^44,45^ This is crucial for the field of transplantation as psychosocial characteristics are often important considerations for candidacy. Furthermore, it is important to note that patients registered in the SRTR were those who have already been placed on the waitlist and its demographic distribution may be a reflection of disparate care that are present in settings more upstream of the care pathway (e.g., specialty referral).^3,46–49^ This may explain why the AI-SC unexpectedly slightly disadvantaged patients from less SES deprived areas, White, and Asian races for transplant. Critical evaluations of bias in LLMs and the field of transplantation will be more important than ever prior to clinical deployment.

In addition to considerations of bias, fine-tuning the AI-SC also revealed the significance of measured clinical endpoints in LT. Model performance was highly dependent on the definition of “success” in transplantation. Since it is emphasized in quality metrics and public perception, this present study defined “success” as 1-year post-LT survival. There is a subtle, yet important difference between the distinction of “unlikely to benefit” and “unlikely to benefit *enough*” to justify use of a scarce resource.^7^ Organizations such as Trillium Gift of Life Network, a provincial organ and tissue donation agency in Canada, have established that patients should not be considered for LT if the likelihood of 5-year post-LT survival is less than 60% though this endpoint can be debated.^50^ Beyond death and graft survival, there is wide variability in post-operative morbidity among centers and relevant clinical endpoints may differ across the spectrum of liver diseases.^51,52^ Patients may also have different definitions of success, stating that restoration of physical function, relief from worry, and ability to meaningfully contribute to society are more valuable metrics, while post-LT complications were a “part of the process”.^53^ While attempting to predict LT benefit based on 1-year post-operative survival is an appropriate initial step, other modifiable post-LT variables that play an important role in a “successful” LT warrant investigation.

How AI adheres to ethical values such as beneficence, utility, and justice will depend on how AI is developed, integrated, and deployed in practice. When surveyed on the beliefs and attitudes towards AI in healthcare, patients still place their trust in providers rather than AI tools.^54^ The potential benefits of AI in speed, efficiency, and reducing administrative workload must be balanced with its limitations in handling nuance, transparency, and explainability.

Among surveyed transplant providers, there is optimism that AI could be a beneficial clinical decision support tool, though its programming and heuristics require scrutiny.^55^ However, our results and discussion should not be interpreted that the AI-SC should be the sole determinant for LT candidacy. When using ML models to risk stratify or estimate survival benefit based on clinical data, it is important to recognize that these data (e.g., patient referral, lab results, encounters) were based on clinically-manifested actions by a human practitioner. The sheer act of gathering and reporting data may bias the LLMs towards outcomes of interest or clinical presuppositions. Baseline performance levels should be exceeded prior to claims that ML models can provide tangible guidance to clinicians. Thus, AI should not replace or supplant the committee as a final decision-maker, rather its applications may lie in its ability to augment fact-finding, gather data, and as safety guardrails while “looking over the shoulders” of members of the LT-SC.^56^

### Limitations

This study has several limitations. Like all large datasets, missing values in the SRTR limited the number of available inputs for analysis, which also restricted the variety of specialists that can be included in the AI-SC. Future work using more variables and long-context clinical notes, leveraging the full strength of ML models to incorporate big data, may reveal more nuanced perspectives among AI specialists.^15,57^ Next, the SRTR only includes patients who were accepted for waitlisting and representative cases of patients with contraindications to LT were synthetic.^26^ Future investigations with prospective cohorts prior to listing would strengthen the applicability and generalizability of the AI-SC. Lastly, the cost of LLMs in this study restricted the size of the data set included in the analysis. Moreover, studies on deployment of AI-SC are still necessary for performance and validation. AI-SCs for LT may be trained to account for center-specific criteria/performance metrics, different subpopulations, and shifting guidelines on previously taboo scenarios (e.g., alcoholic hepatitis, colorectal liver metastasis). How AI decision support tools are woven with clinician judgment is an important future area of investigation. Nevertheless, the use of AI agents in our study is a provocative step towards objectifying the LT selection process, increasing consistency, and relieving the burden placed on LT-SC when making difficult, life and death decisions.

## Conclusions

AI agents can be used to simulate the LT-SC and apply medical domain knowledge of various subspecialists to objectively identify patients who may benefit from LT. The AI-SC’s accuracy for identifying contraindications to LT, predict 6-month, and 1-year survival were 98.2%, 94.9%, and 92.0%, respectively. Future research investigating risks of bias, other clinical endpoints of LT, and the utility of clinical decision support tools in the LT-SC will be critical towards advancing the use of LLMs in transplantation.

## Data Availability

The data that support the findings of this study are publicly available through the Scientific Registry of Transplant Recipients. Its study protocol, and statistical analysis plan will be made available following publication upon request. Please contact the corresponding author via email with a study proposal (e.g., statistical plan, study protocol) and details regarding how the data will be used.

## Acknowledgements

We acknowledge the support of the project from our patient partners, the Transplant AI initiative, and University Health Network (UHN) Foundation.

## Notes

### Competing Interest Statement

The authors have declared no competing interest.

### Funding Statement

This research was supported by funding from the Transplant AI initiative, Ajmera Transplant Centre, University Health Network.

### Author Declarations

This study was approved by the Institutional Review Board at University of California, Irvine (#4474).

