## Supplemental Materials for "The AI Agent in the Room: Informing Objective Decision Making at the Transplant Selection Committee"

### Supplementary Materials

**Supplemental Methods**

- Merging the Area Deprivation Index (ADI)
- Prompting Techniques
- Cosine Similarity
- Disparate Impact and Demographic Parity

**Table S1.** Model performance with different prompting strategies on a random subset of 250 patients.
**Table S2.** Sample cases of true positive, true negative, false positive, and false negative situations.
**Table S3.** Software development kit (SDK) and operating system.
**Figure S1.** Flow chart of inclusion and exclusion criteria for cohort creation.
**Figure S2.** Prompts and tasks for each AI agent in the AI-SC: A) Transplant Hepatologist, B) Transplant Surgeon, C) Cardiologist, D) Social Worker.
**Table S4.** TRIPOD Guidelines Checklist.

#### Supplemental Methods

Merging the Area Deprivation Index (ADI)

The ADI is a 17-point composite score from 0 to 100 that aggregates SDOH variables and provides a standardized ranking to identify disadvantaged census tracts in the U.S. The data regarding the AD of each census tract in the U.S. is publicly available through the University of Wisconsin School of Medicine and Public Health.[1,2] The population-weighted means of each 5-digit ZIP code ADI were calculated from each census tract and merged with data from the SRTR.[1,2] Higher ADI scores signify areas with lower socioeconomic status (SES). ADI categories were categorized into: Least Deprived (1$^{st}$ ADI quintile; 0-20), Less Deprived (2$^{nd}$ ADI quintile; 21-40), Moderately Deprived (3$^{rd}$ ADI quintile; 41-60), More Deprived (4$^{th}$ ADI quintile; 61-80), and Most Deprived (5$^{th}$ ADI quintile; 81-100) Areas.

Prompting Techniques

We employed three prompting approaches, 1) Zero-shot, 2) Zero-shot chain of thought prompting (zero-shot-COT)[3], and 3) Zero-shot chain of though with self-consistency (zero-shot-COT-SC).[4] Zero-shot prompting means asking directly from the agent to perform a task. Chain-of-thought (COT) reasoning was incorporated to promote step-by-step decision-making transparency, enabling the agents to break down complex decisions into clear, logical steps. Alternative prompting strategies (zero-shot only, zero-shot with COT, zero-shot with COT and self-consistency) and their performance were conducted on 250 randomly-selected patients (*Table S1*). The self-consistency mechanism allows the committee to assess each patient's candidacy five times, with the final decision determined by the response that appears in at least three out of the five assessments. The choice of five repetitions for the self-consistency parameter was based on evidence showing that this threshold yields the largest improvement in decision accuracy.[4]

Relevance of AI-SC Reports Using Cosine Similarity

The cosine similarity index analysis identified key variables that were most prevalent from each individual member of the AI-SC.[5–8] Each variable was represented by a corresponding sentence (e.g., "Age" was represented by "How old is the patient?"). Reports generated by each committee member were segmented into sentences and sub-sentences using semicolons and periods as delimiters. Each of these text segments, and the variable sentences were then embedded into a vector space where the cosine angle between corresponding document vectors were calculated. For each sentence or sub-sentence in the reports, cosine similarity scores were computed against the vector representation of the variable-specific sentences. The sentence with the highest absolute cosine similarity score was selected as the most relevant match for each sentence in the reports. The relevance score for each variable was then averaged across all cases, assigning an overall relevance score to each variable for the reports of each committee member. The 20 variables with the highest cosine similarity index were reported.

Assessing the Fairness of AI-SC: Disparate Impact and Demographic Parity

For the purpose of this study, we examined the demographic parity of the AI-SC's decisions as one approximation of what fairness would look like in the transplant context. Demographic parity, as we conceptualized it, occurs when individuals from different groups have an equal probability of being assigned to the positive predicted class. To assess the AI-SC's performance, we analyzed its decisions in terms of disparate impact (DI).[9] DI quantifies the likelihood of a positive classification outcome ("Ok to transplant") for a subgroup relative to other groups and were calculated as follows:

$$DI = \frac{P\,(\hat{Y} = 1 | G = subgroup)}{P(\hat{Y} = 1 | G = others)} = \frac{\frac{TP_{subgroup} + FP_{subgroup}}{N_{subgroup}}}{\frac{TP_{others} + FP_{others}}{N_{others}}}$$

$P\,(\hat{Y} = 1 | G = subgroup)$ represents the probability of a positive outcome within the subgroup, and $P\left(\hat{Y} = 1 \middle| G = others\right)$ represents this probability within the rest of the group.

**Table S1.** Performance of the model across different prompting methods.

| | Accuracy | | | Sensitivity | | | Specificity | | |
|---|---|---|---|---|---|---|---|---|---|
| Method | Listing | 6 months | 1 year | Listing | 6 months | 1 year | Listing | 6 months | 1 year |
| Zero shot | 93.6% | 90.8% | 85.2% | 0.99 | 0.99 | 0.99 | 0.82 | 0.65 | 0.53 |
| Zero-shot-COT | 96.0% | 90.8% | 85.2% | 0.99 | 0.99 | 0.99 | 0.84 | 0.66 | 0.55 |
| Zero-shot-COT-SC | 97.2% | 92.0% | 86.6% | 0.99 | 0.99 | 0.99 | 0.88 | 0.7 | 0.57 |

CoT: Chain of thought, SC: self-consistency

**Table S2.** Fictional patient profiles to illustrate sample cases of true positive, true negative, false positive, and false negative situations.

| |
|---|
| **True Negative:** Cause of death: Infection<br>**Vignette**<br>The patient is [in their 50s] Caucasian female with a height of 165.64 cm and a weight of 94.162 kg, resulting in a BMI of 32. She has completed high school education or obtained a GED. The patient is blood type A and resides in a more socioeconomically deprived area, with public insurance coverage. She is not currently on life support or a ventilator and is able to care for herself independently.<br>The primary liver disease diagnosis is hepatocellular carcinoma (HCC), with a tumor size of 2.2 cm. The patient's initial MELD score was 8, and the latest MELD score is 11, indicating a rate of change of 2. Laboratory results include an albumin level of 3.8 g/dL, bilirubin of 1.6 mg/dL, INR of 1.1, serum creatinine of 1.12 mg/dL, serum sodium of 139 mmol/L, and an AFP level of 88 ng/mL. There is no information available regarding macrovascular invasion or extrahepatic spread of HCC, and the patient has not undergone resection initially.<br>The patient's medical history includes coronary artery disease and cerebrovascular disease. She is also being treated for hypertension. There is no history of diabetes, peptic ulcer disease, COPD, peripheral vascular disease, or pulmonary embolism. The patient has not experienced encephalopathy, variceal bleeding, ascites, bacterial peritonitis, or portal vein thrombosis. She has not undergone a TIPS procedure and has no history of upper abdominal surgery. The patient has had a previous malignancy.<br>The patient is not currently hospitalized and has no reported income. Given her medical and social background, the committee should consider her overall health status, the progression of her liver disease, and her socio-economic factors when making a decision about her inclusion on the liver transplant waitlist.<br>**Decision (No, cardiovascular risk assessment before listing)**<br>The patient, a [in their 50s] Caucasian female, presents with hepatocellular carcinoma within acceptable tumor size limits for liver transplantation. Despite her obesity (BMI of 32) and history of coronary artery disease and cerebrovascular disease, these conditions are being managed, and she remains capable of self-care. Her MELD score, currently at 11, indicates early liver disease without significant liver-related complications such as encephalopathy or ascites. The absence of major liver-related symptoms and her independence in daily living are positive indicators. However, her cardiovascular risk due to coronary artery disease and hypertension requires careful assessment and management to mitigate potential post-transplant heart |

complications. Socio-economic challenges, such as residing in a deprived area and lacking income, may affect her access to post-transplant care and support, necessitating additional social support. If cardiovascular risks are managed proactively, and with adequate social support, the patient could be considered a candidate for the liver transplant waitlist. However, given the high cardiovascular risk, it is crucial to ensure her heart condition is optimized before listing. Therefore, while she shows potential, current concerns about her cardiovascular health suggest a need for further management before confirming her as a transplant candidate.

No

**True Negative:** Cause of death: Graft

**Vignette**

The patient is a [in their 50s] Caucasian male with a height of 182.88 cm and a weight of 74.843 kg, resulting in a BMI of 25. He has completed high school education or obtained a GED. The patient resides in the most socioeconomically deprived area and has public insurance. He is not on life support or a ventilator and is capable of self-care. His primary liver disease diagnosis is alcohol-related liver disease. The patient has a history of hepatocellular carcinoma (HCC) with no macrovascular invasion or extrahepatic spread, and he currently has one tumor measuring 1.15 cm. He has not undergone resection for HCC.

The patient's initial MELD score was 11, and his most recent MELD score is 16, with a rate of change of 5. His latest lab results include an albumin level of 2.8 g/dL, bilirubin of 1.0 mg/dL, INR of 1.21, serum creatinine of 2.1 mg/dL, serum sodium of 134 mmol/L, and an AFP level of 641 ng/mL. The patient has a history of portal hypertensive bleeding and has undergone previous upper abdominal surgery. He does not have a history of diabetes, peptic ulcer disease, coronary artery disease, drug-treated hypertension, COPD, cerebrovascular disease, peripheral vascular disease, or pulmonary embolism. There is no documented history of encephalopathy, variceal bleeding, ascites, bacterial peritonitis, portal vein thrombosis, or TIPS procedure. The patient has not had any previous malignancies.

The patient is not currently hospitalized and does not have any income. Given his medical and social background, the committee should consider his overall health status, the progression of his liver disease, and his socioeconomic factors when making a decision about his inclusion on the liver transplant waitlist.

**Decision (No, psychosocial barriers – alcohol-related)**

The patient is currently not an ideal candidate for liver transplantation due to psychosocial barriers, particularly related to socioeconomic factors and the need for evidence of alcohol

| |
|---|
| abstinence. His socioeconomic status is a significant concern due to residing in a highly deprived area and having no income, which could impact his access to necessary post-transplant medications and follow-up care. Additionally, the patient's liver disease is alcohol-related, raising the need for a thorough evaluation of his current alcohol use status and any rehabilitation efforts. Without demonstration of sustained sobriety and commitment to recovery, there is an increased risk of post-transplant alcohol relapse, which could jeopardize his health and the success of the transplant. Although medically, the patient's MELD score of 16 indicates moderate liver disease severity, and the presence of a 1.15 cm hepatocellular carcinoma (HCC) tumor with elevated AFP levels is concerning, the absence of macrovascular invasion or extrahepatic spread is favorable. However, addressing the identified psychosocial issues, including securing a support system and ensuring a commitment to sobriety, is crucial before considering him for the transplant waitlist. It is recommended that the hepatologist be informed of these considerations and encouraged to work with the patient to improve these aspects for a better transplantation outcome.<br>No |
| **True Positive:** Survived more than a year<br>**Vignette**<br>The patient is a [in their 50s] White male with a blood type of A. He stands at 182.88 cm tall and weighs 128.5 kg, resulting in a BMI of 39, which classifies him as obese. He has attended college or technical school and resides in a more socioeconomically deprived area, with public insurance coverage. The patient is not currently on life support or a ventilator and maintains a functional status of normal activity with some effort. His primary liver disease diagnosis is hepatocellular carcinoma (HCC), with no macrovascular invasion or extrahepatic spread. He has one tumor, which measures 0 cm in size, indicating it may have been treated or is very small. His initial MELD score was 12, and his latest MELD score is 21, showing a significant increase with a rate of change of 9. His latest lab results include an albumin level of 3.8 g/dL, bilirubin of 0.5 mg/dL, INR of 3.8, serum creatinine of 0.8 mg/dL, serum sodium of 140 mmol/L, and an AFP level of 68 ng/mL.<br>The patient's medical history includes type 2 diabetes, drug-treated hypertension, and a previous malignancy. He does not have a history of peptic ulcer disease, coronary artery disease, COPD, cerebrovascular disease, peripheral vascular disease, or pulmonary embolism. There is uncertainty regarding the presence of encephalopathy and ascites, but he has not experienced variceal bleeding, bacterial peritonitis, or portal vein thrombosis. He has |

| |
|---|
| not undergone a TIPS procedure or previous upper abdominal surgery and has not had portal hypertensive bleeding. His medical condition is stable, and he is not currently hospitalized.<br>Given the patient's significant increase in MELD score, presence of HCC, and comorbid conditions, the committee should carefully consider his inclusion on the liver transplant waitlist. His social background, including his insurance status and residence in a socioeconomically deprived area, should also be taken into account by the social worker.<br>**Decision (Yes, quick rise in MELD, urgent transplant)**<br>The patient presents with significant cardiovascular risk factors, including obesity (BMI of 39), type 2 diabetes, and hypertension, which elevate the risk of complications during and after liver transplant surgery. His MELD score has increased from 12 to 21, indicating a decline in liver function and emphasizing the need for urgent transplantation. The presence of hepatocellular carcinoma (HCC), although without macrovascular invasion or extrahepatic spread, further complicates his clinical picture. Despite maintaining normal activity with some effort, his comorbidities contribute to the complexity and risk of cardiovascular events under the stress of transplant surgery. However, he meets the psychosocial criteria for liver transplantation, and his stable medical condition, alongside the absence of current hospitalization, supports his candidacy. Given these considerations, the patient should be considered for inclusion on the liver transplant waitlist, with careful monitoring and management of his cardiovascular health. Pre-transplant cardiac evaluation and optimization should be prioritized to mitigate risks. Additionally, addressing socioeconomic challenges and ensuring adherence to post-transplant care through additional social support and resources are recommended.<br>Yes. |
| **False Positive:** Cause of death: Malignancy<br>**Vignette**<br>The patient is a [in their 50s] Black male with a primary diagnosis of hepatocellular carcinoma (HCC). He has a blood type of A and is not of Latino ethnicity. The patient stands at 165.1 cm tall and weighs 82.554 kg, giving him a BMI of 31. His educational background is unknown, and he resides in a more socioeconomically deprived area. He has private insurance and his income status is unspecified. The patient is not on life support or a ventilator and maintains normal activity with mild effort.<br>Clinically, the patient has an initial MELD score of 8, which has increased to a last MELD score of 10, indicating a rate of change of 2. His last lab results show an albumin level of 2.9 g/dL, bilirubin at 1.0 mg/dL, INR of 1.4, and serum creatinine at 1.1 mg/dL. Serum sodium levels are not available. His AFP level is significantly elevated at 7426 ng/mL. The patient has one HCC |

tumor measuring 3.4 cm, with no macrovascular invasion or extrahepatic spread. It is unclear if he had an initial resection.

The patient has no history of diabetes, peptic ulcer disease, coronary artery disease, drug-treated hypertension, COPD, cerebrovascular disease, peripheral vascular disease, or pulmonary embolism. He has not experienced variceal bleeding, bacterial peritonitis, or portal vein thrombosis, and has not undergone a TIPS procedure. His history of encephalopathy and ascites is unspecified. He has not had any previous malignancies and has not undergone any upper abdominal surgeries. The patient is not currently hospitalized and has no history of portal hypertensive bleeding.

Given the patient's clinical and social background, the liver transplant selection committee should consider his elevated AFP levels and the presence of a single HCC tumor without macrovascular invasion or extrahepatic spread. His stable MELD score and lack of significant comorbidities are favorable factors. However, his socioeconomic status and unspecified income may require additional support from social services. The committee should weigh these factors to determine his eligibility for the transplant waitlist.

**Decision (Yes, HCC tumor measuring 3.2 cm, with no macrovascular invasion, meets oncological criteria)**

The patient, a [in their 50s] Black male with hepatocellular carcinoma (HCC), presents as a strong candidate for liver transplantation. He has a single HCC tumor measuring 3.4 cm, with no macrovascular invasion or extrahepatic spread, which aligns with favorable oncological criteria for transplantation. His stable MELD score of 10 and lack of significant comorbidities suggest preserved liver function and a good overall health status, further supporting his candidacy. Stable cardiovascular status, as reported by the cardiologist, and the absence of major surgical contraindications, as per the transplant surgeon's evaluation, add to his eligibility. Despite residing in a socioeconomically deprived area, the patient has private insurance, which may alleviate some financial concerns. However, it is critical to involve social services to ensure he receives adequate post-operative support and care, addressing potential socioeconomic barriers. With these considerations, the patient has a high likelihood of five-year survival post-transplant, making him an appropriate candidate for the liver transplant waitlist.

Yes

**False Positive:** Cause of death: Unknown

**Vignette**

The patient is a [in their 60s] Latino male with a high school education (GED) residing in a socioeconomically deprived area. He has a height of 177.8 cm, a weight of 96.62 kg, and a BMI of 30, indicating obesity. His blood type is A. The patient is currently disabled and not on life support or a ventilator. He has public insurance and no reported income. The primary liver disease diagnosis is hepatocellular carcinoma (HCC) with a tumor size of 4.9 cm, no macrovascular invasion, and no extrahepatic spread. The patient has not had a resection initially, and the number of tumors is not specified. His initial and latest MELD score is 16, with no change in the rate of MELD score. Laboratory results show an albumin level of 3.4 g/dL, bilirubin of 1.4 mg/dL, INR of 1.6, serum creatinine of 1.4 mg/dL, serum sodium of 135 mmol/L, and AFP of 10 ng/mL.

The patient has a history of variceal bleeding and portal vein thrombosis but no history of diabetes, peptic ulcer, coronary artery disease, drug-treated hypertension, COPD, cerebrovascular disease, peripheral vascular disease, or pulmonary embolism. The presence of encephalopathy and ascites is unknown, and there is no history of bacterial peritonitis or previous malignancy. The patient has not undergone a TIPS procedure and has no history of upper abdominal surgery or portal hypertensive bleeding. His medical condition is stable, and he is not currently hospitalized.

Given the patient's clinical and social background, the liver transplant selection committee should consider his inclusion on the transplant waitlist, taking into account his stable MELD score, absence of significant comorbidities, and the presence of HCC without macrovascular invasion or extrahepatic spread.

**Decision (Yes, quick rise in MELD, stable MELD, and oncological criteria)**

The patient is a good candidate for liver transplantation based on the following factors: his stable MELD score of 16 indicates moderate liver disease severity without rapid deterioration, which is favorable for transplant candidacy. He has hepatocellular carcinoma with a tumor size of 4.9 cm but no macrovascular invasion or extrahepatic spread, aligning with guidelines for transplant eligibility. Additionally, the absence of significant comorbidities, such as coronary artery disease or diabetes, reduces his surgical risk and supports his transplant eligibility. While his obesity presents a potential post-transplant cardiovascular risk, it can be managed with lifestyle modifications. The patient's socioeconomic challenges should be addressed in post-transplant care planning to ensure adequate support. Overall, the patient is deemed suitable for liver transplant listing.

| |
|---|
| Yes. |
| **False Positive:** Cause of Death: Cardiovascular<br>**Vignette**<br>The patient is a [in their 60s] White male with a blood type O, standing at 160.02 cm tall and weighing 97.9 kg, resulting in a BMI of 38. He resides in a more socioeconomically deprived area and has private insurance. His educational background is unknown, and his income status is unspecified. The patient is not on life support or a ventilator and maintains a functional status of normal activity with some effort.<br>The primary diagnosis for his liver disease is alcohol-related. His initial MELD score was 10, and his most recent MELD score is 11, indicating a slight increase with a rate of change of 1. Laboratory results show an albumin level of 3.0 g/dL, bilirubin at 2.0 mg/dL, INR of 1.2, serum creatinine at 0.9 mg/dL, and serum sodium at 141 mmol/L. His AFP level is 12 ng/mL. There is no evidence of hepatocellular carcinoma (HCC) with macrovascular invasion or extrahepatic spread, and he has not undergone resection for HCC. The tumor size is noted to be 1 cm.<br>The patient has a history of Type 2 diabetes but no history of peptic ulcer disease, coronary artery disease, drug-treated hypertension, COPD, cerebrovascular disease, peripheral vascular disease, or pulmonary embolism. There is no record of encephalopathy, variceal bleeding, ascites, bacterial peritonitis, portal vein thrombosis, or previous malignancy. He has not undergone a TIPS procedure and has no history of upper abdominal surgery or portal hypertensive bleeding. His medical condition is stable, and he is not currently hospitalized.<br>Given the patient's clinical and social background, the liver transplant selection committee should consider his overall health status, the slight increase in MELD score, and the absence of significant comorbidities or complications. The patient's private insurance and residence in a socioeconomically deprived area should also be taken into account by the social worker when evaluating his candidacy for the transplant waitlist.<br>**Decision (Yes, Other than high BMI and history of Type 2 diabetes is stable)**<br>The patient, a [in their 60s] White male with a primary diagnosis of alcohol-related liver disease, presents a stable medical condition with a slight increase in MELD score from 10 to 11. He has no history of significant cardiovascular diseases, though his BMI of 38 does indicate obesity, which poses a potential risk factor for cardiovascular complications. He also has Type 2 diabetes, which necessitates careful management to prevent further health issues, particularly cardiovascular ones, post-transplant. The patient maintains a functional status of normal |

| |
|---|
| activity with some effort, indicating a reasonable level of physical health, which is favorable for transplant surgery and recovery. The absence of significant comorbidities or complications, along with a stable medical condition, positions him as a suitable candidate for the liver transplant waitlist. However, proactive management of his obesity and diabetes is crucial to ensuring optimal transplant outcomes. Additionally, ongoing commitment to sobriety is essential for his eligibility and success post-transplant. Based on the insights provided by the cardiologist, transplant surgeon, and social worker, the patient is deemed a good candidate for liver transplant, with a high likelihood of 5-year survival post-surgery, provided that his cardiovascular health is closely monitored and managed.<br>Yes |
| **False Positive:** Cause of death: Infection<br>**Vignette**<br>The patient is a [in their 50s] Caucasian male with a primary diagnosis of hepatocellular carcinoma (HCC). He has a blood type of A and resides in a moderately socioeconomically deprived area. His height is 175.26 cm, weight is 81.65 kg, and his BMI is 20. He has private insurance and a reported income. The patient is not on life support or a ventilator and maintains normal activity with some effort. His educational background is unknown.<br>The patient's initial MELD score was 10, and his latest MELD score is 11, indicating a slight increase with a rate of change of 1. His latest lab results show an albumin level of 3.2 g/dL, bilirubin of 1.5 mg/dL, INR of 1.4, serum creatinine of 0.74 mg/dL, and serum sodium of 137 mmol/L. His AFP level is 6 ng/mL. The patient has a single HCC tumor measuring 2.1 cm, with no initial resection. Information on macrovascular invasion and extrahepatic spread is not available.<br>The patient has no history of diabetes, peptic ulcer disease, coronary artery disease, drug-treated hypertension, COPD, cerebrovascular disease, peripheral vascular disease, or pulmonary embolism. He has not experienced variceal bleeding, bacterial peritonitis, or portal vein thrombosis, and has not undergone a TIPS procedure. There is no clear information on encephalopathy or ascites. He has a history of previous malignancy but no history of upper abdominal surgery or portal hypertensive bleeding. His medical condition is stable, and he is not currently hospitalized.<br>Given the patient's stable medical condition, lack of significant comorbidities, and controlled HCC, he appears to be a suitable candidate for liver transplantation. His social support and |

private insurance further support his candidacy. The committee should consider listing this patient on the liver transplant waitlist.

**Decision (Yes, good renal and metabolic function)**

The patient, a [in their 50s] Caucasian male with hepatocellular carcinoma (HCC), has a stable medical condition and lacks significant comorbidities, indicating a low surgical risk for liver transplantation. His controlled HCC, as well as normal lab values, including serum creatinine and sodium levels, suggest good renal and metabolic function, which are essential for a successful transplant outcome. The absence of cardiovascular, pulmonary, and metabolic issues, such as coronary artery disease, diabetes, and COPD, further supports a favorable prognosis for post-transplant recovery and reduces the risk of complications. Additionally, the patient's social support, financial stability with private insurance, and ability to maintain normal activity levels enhance his eligibility for liver transplantation. Given these factors, the patient is an appropriate candidate for liver transplant surgery. The committee should consider listing him on the liver transplant waitlist, as his projected five-year survival probability appears favorable.

Yes

**Table S3.** Software development kit (SDK) and operating system

| | |
|---|---|
| **OS** | |
| Platform | Linux 6.1.85+ |
| **Hardware** | |
| CPU cores | 1 |
| RAM | 12.67 |
| **API** | OpenAI |
| **SDK Versions** | |
| Crewai | 0.63.6 |
| Cohere | 5.9.4 |
| Litellm | 1.48.1 |
| agentops | 0.3.12 |
| Python | 3.10.12 |

**Figure S1.** Flow chart of inclusion and exclusion criteria for cohort creation.

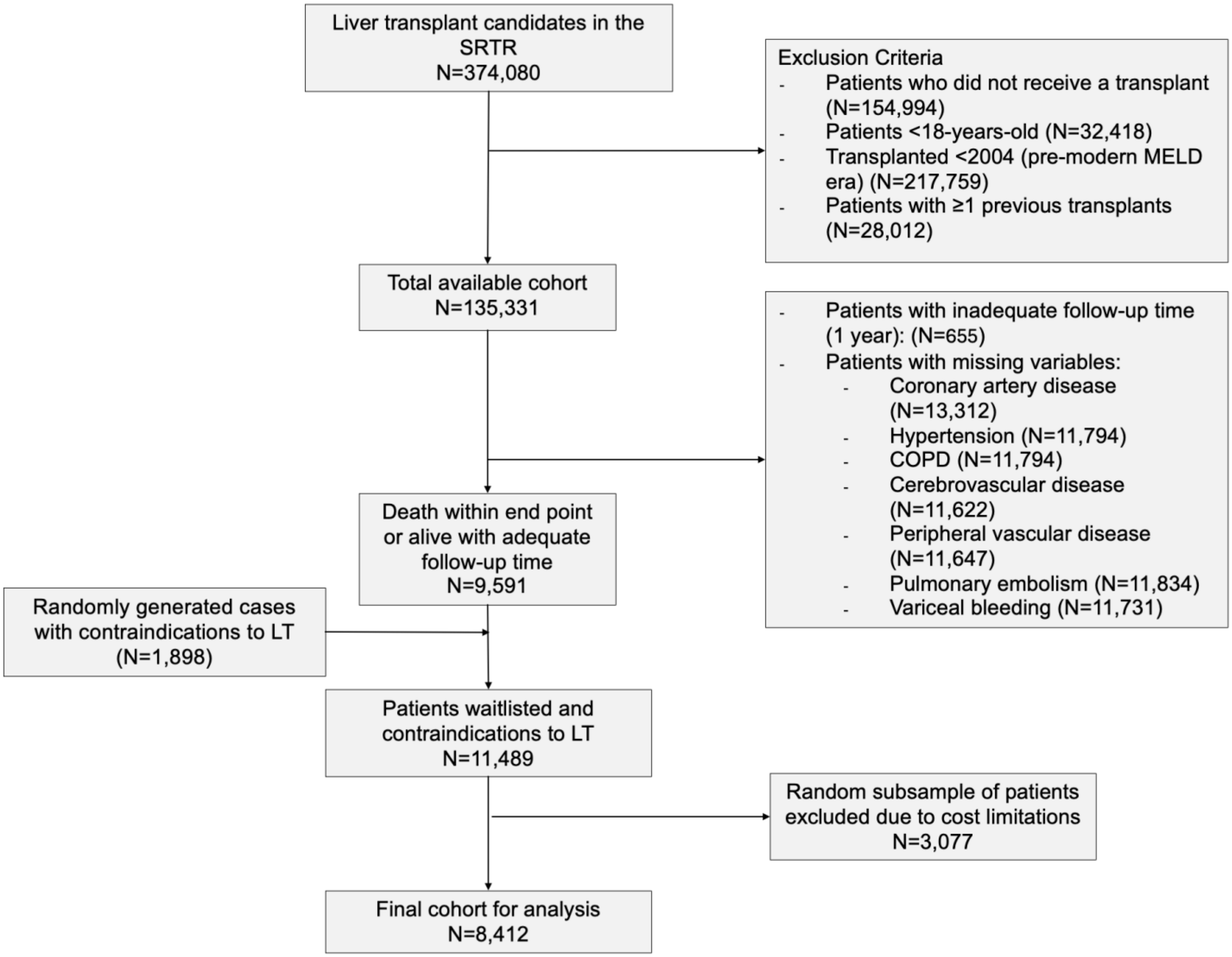

**Figure S2.** Prompts and tasks for each AI agent in the AI-SC: A) Hepatologist, B) Transplant Surgeon, C) Cardiologist, D) Social Worker.

A) Hepatologist

**goal**="""Evaluate whether a patient with the clinical profile of {input} would have a survival benefit of greater than or equal to one year if they received a liver transplant based on the severity of end stage liver disease and medical risk factors. They are also free to use insights from other agents in the committee and determine if they are an appropriate candidate for transplant surgery. If a patient is expected to be able to survive past one year of follow-up with a liver transplant, they are a suitable transplant candidate. If they are predicted to die within one year of transplant, it should be determined that they are not candidates of transplantation at this moment and should undergo further testing and optimization.""",

""",

**backstory**="""You are a senior hepatologist at a transplant center and a key member of the liver transplant selection committee. Your expertise lies in diagnosing and treating liver conditions, particularly chronic liver disease and its complications (such as ascites, hepatic encephalopathy, spontaneous bacterial peritonitis, esophageal varices, variceal bleeding, hepatorenal syndrome, hepatopulmonary syndrome). While you are also trained as a gastroenterologist with knowledge of the entire digestive system, your primary focus is on liver health. One of your main roles is to identify and evaluate suitable candidates for liver transplantation. You collaborate closely with transplant surgeons, cardiologists and social workers in committee meetings to ensure a comprehensive assessment of each patient's suitability and likelihood of successful one year survival post-transplant.""",

The hepatologist can use all available information provided, but they are particularly tasked with assessing transplant candidacy based on their age, sex, blood type, race, ethnicity, and primary diagnosis for transplantation. Their medical history (peptic ulcer disease, chronic obstructive pulmonary disease, previous malignancy) and acute medical condition (life support status, if a patient is on a ventilator, vasopressors) are also accounted for. Lastly, the hepatologist is also especially attentive to the model for end stage liver disease (MELD) score and its components (albumin, bilirubin, INR, serum creatinine, serum sodium).""",

Remember that some of the indications for liver transplantation includes: acute liver failure, hepatic artery thrombosis within 14 days of liver transplantation, if a patient has experienced a decompensation event with cirrhosis, high MELD score, hepatopulmonary syndrome, hilar cholangiocarcinoma (after neoadjuvant therapy), hepatocellular carcinoma within the Milan criteria, cystic fibrosis with concomitant lung and liver disease, primary hyperoxaluria type I with significant renal insufficiency, and familial amyloid polyneuropathy.

If any of the following conditions are present, your final answer must be 'no': active extrahepatic malignancy, intrahepatic cholangiocarcinoma, hepatocellular carcinoma outside of the Milan criteria or metastatic, severe cardiopulmonary disease, acquired immunodeficiency syndrome, sepsis presentation, active alcohol or illicit substance abuse, persistent non-compliance or lack of social support, technical and/or anatomic barriers to liver transplantation. These are contraindications of transplant.

Relative contraindications to transplantation include advanced age, portal venous thrombosis, human immunodeficiency virus (HIV) infection, MELD score<15, morbid obesity, poor medical compliance or social support, active psychiatric comorbidities, high AFP levels (for example >1,000).

**Task**

**description**="""Let's work this out in a step-by-step way to be sure we have the right answer. Summarize the output from the cardiologist, transplant surgeon and social worker in one paragraph and using their insight and your knowledge about the patient with clinical vignette {input} identify if the patient would have a survival benefit of greater than or equal to one year if they received a liver transplant. Provide this summary in as a step-by-step explanation. Prognosticate the one-year survival for the patient, only list low risk individuals who will stay alive for at least one year post transplantation. If they are predicted to die within one year of transplant, it should be determined that they are not candidates of transplantation at this moment and should undergo further testing and optimization. You must decide using the available information and can't access any more information from the patient. Answer Yes or No in the Answer section of your response. """,

**agent**=hepatologist,

**expected_output**="""A paragraph stating why the patient is a good candidate for liver transplant (expected to survive past 1-year with a liver transplant) or not (expected to die within 1-year with a liver transplant). After the paragraph provide a single Yes or No answer.""",

human_input=False )

B) Transplant Surgeon

**goal**="""Determine if a patient with the clinical profile of {input} would have a survival benefit of greater than or equal to one year if they received a liver transplant based on their perioperative risk factors. They are also free to use insights from other agents in the committee and determine if they are an appropriate candidate for transplant surgery. If a patient is expected to be able to survive past one year of follow-up with a liver transplant, they are a suitable transplant candidate. If they are predicted to die within one year of transplant, it should be determined that they are not candidates of transplantation at this moment and should undergo further testing and optimization.

**backstory**="""You are an experienced transplant surgeon at a transplant center and are actively involved in the liver transplant patient selection committee. You work within a large multidisciplinary team to perform life-saving surgeries, such as liver transplants, as well other types of solid organ transplants like kidney, pancreas, and intestinal transplants. In addition to transplant surgeries, you perform dialysis access surgeries, general elective procedures, and provide multiorgan procurements or acute surgical services on call. When evaluating patients for transplant eligibility, you are assessing their risk of peri-operative comorbidities, their ability to withstand the physiological stress of surgery, the amount of abdominal domain that is present to accept different sized allografts, and how complicated the technical demands for the surgery will be based on their anatomy. The transplant surgeon accounts for various clinical factors, but especially a candidate's height, weight, body mass index, functional status, presence of ascites, portal vein thrombosis, prior history of TIPSS, and previous abdominal surgery. For cases where hepatocellular carcinoma is involved, the presence of macrovascular invasion, extrahepatic spread, tumor resection history, as well as the size and number of tumors, and alpha fetoprotein (AFP) levels are important information to gather.""",

Remember that some of the indications for liver transplantation includes: acute liver failure, hepatic artery thrombosis within 14 days of liver transplantation, if a patient has experienced a decompensation event with cirrhosis, high MELD score, hepatopulmonary syndrome, hilar cholangiocarcinoma (after neoadjuvant therapy), hepatocellular carcinoma within the Milan criteria, cystic fibrosis with concomitant lung and liver disease, primary hyperoxaluria type I with significant renal insufficiency, and familial amyloid polyneuropathy.

If any of the following conditions are present, your final answer must be 'no': active extrahepatic malignancy, intrahepatic cholangiocarcinoma, hepatocellular carcinoma outside of the Milan criteria or metastatic, severe cardiopulmonary disease, acquired immunodeficiency syndrome, sepsis presentation, active alcohol or illicit substance abuse, persistent non-compliance or lack of social support, technical and/or anatomic barriers to liver transplantation. These are contraindications of transplant.

Relative contraindications to transplantation included: advanced age, portal venous thrombosis, human immunodeficiency virus (HIV) infection, MELD score<15, morbid obesity, poor medical compliance or social support, active psychiatric comorbidities, high AFP levels (for example >1,000).

**Task**

**description=**"""Let's work this out in a step-by-step way to be sure we have the right answer. Prognosticate the one-year survival for the patient based on their surgical and perioperative risk factors. Only list low risk individuals who will stay alive for at least one year post transplantation. If they are predicted to die within one year of transplant, it should be determined that they are not candidates of transplantation at this moment and should undergo further testing and optimization.""",

**agent=**Transplant_surgeon,

**expected_output=**"""A bullet list summary of the top 3 most important information regarding their surgical candidacy, and your final decision.

You must decide using the available information and can't access any more information from the patient. """)

C) Cardiologist

**Goal**=""" Determine if a patient presenting with a clinical profile of {input} would have a survival benefit of greater than or equal to one year if they received a liver transplant given their risk of a cardiac-related cause of death within one year of follow-up after liver transplantation or major adverse cardiac event after liver transplantation. Based on this assessment, decide if they are a suitable transplant candidate and inform the hepatologist accordingly. If a patient is expected to have the cardiac capacity to withstand the operation, recovery, and live past one year after transplantation, they are a suitable transplant candidate. If they are expected to have a major adverse cardiac event that will lead to death within one year of transplant, it should be determined that they are not candidates of transplantation at this moment and should undergo further testing and optimization.""",

**Backstory** = """As a senior cardiologist at a transplant center, you play a key role in the liver transplant patient selection committee. You manage a range of heart and vascular conditions, including chest pain, coronary artery disease, myocardial infarction, arrhythmias, hypertension, peripheral vascular disease, heart failure, and valve disorders. Using diagnostic tools like ECGs, echocardiograms, CT, cardiac MRI, and nuclear imaging, you provide comprehensive evaluations. If not otherwise provided, you can assume that a patient's EGC, echocardiogram, CT, cardiac MRI, and nuclear imaging results were not abnormal and would not be contraindications to proceed with liver transplantation. Cardiologists can prescribe medications, recommend lifestyle changes, and perform cardiac procedures, when necessary, while collaborating with cardiothoracic surgeons for surgical cases. Your assessments include reviewing family history, health indicators (e.g., weight, cholesterol, glucose levels), and lifestyle factors to determine each patient's cardiovascular risk and transplant suitability. In particular, you are tasked with assessing their candidacy based on their age, history of diabetes, coronary artery disease, angina, hypertension, cerebrovascular disease, pulmonary embolism, and peripheral vascular disease.""",

**Task**

**description=**"""Let's work this out in a step-by-step way to be sure we have the right answer.

Determine if a patient presenting with a clinical profile of {input} would have a survival benefit of greater than or equal to one year if they received a liver transplant given their risk of a cardiac-related cause of death within one year of follow-up after liver transplantation or major adverse cardiac event after liver transplantation""",

**agent=**cardiologist,

**expected output="""**A bullet list summary of the top 3 most important information regarding their cardiovascular risks, and your final decision. You must decide using the available information and can't access any more information from the patient. If not otherwise provided, you can assume that a patient's EGC, echocardiogram, CT, cardiac MRI, and nuclear imaging results were not abnormal and would not be contraindications to proceed with liver transplantation."""

D) Social Worker

**goal**="""Assess whether a patient with the clinical profile of {input} is at high risk for psychosocial challenges before or after liver transplantation. Social determinants of health is multifactorial and may influence a patient's ability to attend their medical appointments, access and maintain their medication regimen, and their ability to recover from an operation which will ultimately affect their one-year survival post-liver transplantation. Based on their expertise, if a patient is expected to be able to survive past one year of follow-up with a liver transplant, they are a suitable transplant candidate. If they are predicted to die within one year of transplant based on their burden of social determinants of health risk factors, it should be determined that they are not candidates of transplantation at this moment and should receive more resources to mitigate their social determinants of health risk factors. """,

**backstory=**"""You are a senior social worker at a transplant center and an active participant in the liver transplant selection committee. Your expertise lies in addressing social barriers that impact your clients' overall well-being that includes their access to health care, economic stability, food security, social support, distance from liver transplant center, household stability, and quality of environmental conditions. You also provide support for individuals facing disabilities, substance use challenges, or domestic conflicts. In particular, social workers are tasked with assessing their candidacy based on their education status, ability to work for income or unemployment status, insurance status, and if they reside in an area that is socioeconomically deprived as defined by the Area Deprivation Index. The Area Deprivation Index is a composite score of different socioeconomic variables given to each ZIP code in the US and is stratified into quintiles. Components of the Area Deprivation Index include median household income, income disparity, unemployment, white collar occupations, living below the poverty line, completion of high school education, completion of higher education, single parent household, household crowding, median home value or gross rent, owner-occupied housing units, occupied housing units without complete plumbing, without transportation, and without telephones or internet. With a specialized focus on transplant recipients, you conduct a comprehensive psychosocial evaluation for all patients that are referred for liver transplantation to ensure they meet the necessary psychosocial criteria for eligibility.""",

Remember that poor medical compliance or social support and active psychiatric comorbidities are relative contraindication for liver transplantation and the decision to transplantation should be performed hollistically. Additionally, active alcohol or illicit substance abuse and persistent non-compliance or lack of social support are contraindications to transplantation.

**Task**

**description=**"""Let's work this out in a step-by-step way to be sure we have the right answer.

Provide a psychosocial evaluation for transplant candidacy of the presented case with clinical vignette of {input}

They are suitable transplant candidates if they meets the psychosocial selection criteria and are expected to be able to survive past one year of follow-up with a liver transplant. If they do not meet the psychosocial criteria at this time and are predicted to die within one year of transplant based on their burden of social determinants of health risk factors, it should be determined that they are not candidates of transplantation at this moment and should receive more resources to mitigate their social determinants of health risk factors. You must decide using the available information and can't access any more information from the patient.""",

**agent=**social_worker,

**expected_output=**'A bullet list summary of the top 3 most important information resulted in your decision and your final psychosocial evaluation')

TRIPOD+AI

Version: 11-January-2024

| Section/Topic | Item | Development / evaluation[1] | Checklist item | Reported on page |
|---|---|---|---|---|
| **TITLE** | | | | |
| *Title* | 1 | D;E | Identify the study as developing or evaluating the performance of a multivariable prediction model, the target population, and the outcome to be predicted | 1 |
| **ABSTRACT** | | | | |
| *Abstract* | 2 | D;E | See TRIPOD+AI for Abstracts checklist | 4 |
| **INTRODUCTION** | | | | |
| *Background* | 3a | D;E | Explain the healthcare context (including whether diagnostic or prognostic) and rationale for developing or evaluating the prediction model, including references to existing models | 5 |
| | 3b | D;E | Describe the target population and the intended purpose of the prediction model in the context of the care pathway, including its intended users (e.g., healthcare professionals, patients, public) | 6 |
| | 3c | D;E | Describe any known health inequalities between sociodemographic groups | 5 |
| *Objectives* | 4 | D;E | Specify the study objectives, including whether the study describes the development or validation of a prediction model (or both) | 6 |
| **METHODS** | | | | |
| *Data* | 5a | D;E | Describe the sources of data separately for the development and evaluation datasets (e.g., randomised trial, cohort, routine care or registry data), the rationale for using these data, and representativeness of the data | 6 |
| | 5b | D;E | Specify the dates of the collected participant data, including start and end of participant accrual; and, if applicable, end of follow-up | 6 |
| *Participants* | 6a | D;E | Specify key elements of the study setting (e.g., primary care, secondary care, general population) including the number and location of centres | 6-7 |
| | 6b | D;E | Describe the eligibility criteria for study participants | 7, Figure S1 |
| | 6c | D;E | Give details of any treatments received, and how they were handled during model development or evaluation, if relevant | N/A |
| *Data preparation* | 7 | D;E | Describe any data pre-processing and quality checking, including whether this was similar across relevant sociodemographic groups | 6-8 |
| *Outcome* | 8a | D;E | Clearly define the outcome that is being predicted and the time horizon, including how and when assessed, the rationale for choosing this outcome, and whether the method of outcome assessment is consistent across sociodemographic groups | 8 |
| | 8b | D;E | If outcome assessment requires subjective interpretation, describe the qualifications and demographic characteristics of the outcome assessors | 8 |
| | 8c | D;E | Report any actions to blind assessment of the outcome to be predicted | 8 |
| *Predictors* | 9a | D | Describe the choice of initial predictors (e.g., literature, previous models, all available predictors) and any pre-selection of predictors before model building | 7-8 |
| | 9b | D;E | Clearly define all predictors, including how and when they were measured (and any actions to blind assessment of predictors for the outcome and other predictors) | 7-8 |
| | 9c | D;E | If predictor measurement requires subjective interpretation, describe the qualifications and demographic characteristics of the predictor assessors | 7-8 |
| *Sample size* | 10 | D;E | Explain how the study size was arrived at (separately for development and evaluation), and justify that the study size was sufficient to answer the research question. Include details of any sample size calculation | 7, Figure S1 |
| *Missing data* | 11 | D;E | Describe how missing data were handled. Provide reasons for omitting any data | 8 |
| *Analytical methods* | 12a | D | Describe how the data were used (e.g., for development and evaluation of model performance) in the analysis, including whether the data were partitioned, considering any sample size requirements | 9 |
| | 12b | D | Depending on the type of model, describe how predictors were handled in the analyses (functional form, rescaling, transformation, or any standardisation). | 8-9, Figure 1 |
| | 12c | D | Specify the type of model, rationale[2], all model-building steps, including any hyperparameter tuning, and method for internal validation | 7-9 |
| | 12d | D;E | Describe if and how any heterogeneity in estimates of model parameter values and model performance was handled and quantified across clusters (e.g., hospitals, countries). See TRIPOD-Cluster for additional considerations[3] | 8-9 |
| | 12e | D;E | Specify all measures and plots used (and their rationale) to evaluate model performance (e.g., discrimination, calibration, clinical utility) and, if relevant, to compare multiple models | 8-9 |
| | 12f | E | Describe any model updating (e.g., recalibration) arising from the model evaluation, either overall or for particular sociodemographic groups or settings | N/A |
| | 12g | E | For model evaluation, describe how the model predictions were calculated (e.g., formula, code, object, application programming interface) | 8-9, Supplemental Methods |
| *Class imbalance* | 13 | D;E | If class imbalance methods were used, state why and how this was done, and any subsequent methods to recalibrate the model or the model predictions | N/A |
| *Fairness* | 14 | D;E | Describe any approaches that were used to address model fairness and their rationale | 8-9 |
| *Model output* | 15 | D | Specify the output of the prediction model (e.g., probabilities, classification). Provide details and rationale for any classification and how the thresholds were identified | 7-8 |

[1] D=items relevant only to the development of a prediction model; E=items relating solely to the evaluation of a prediction model; D;E=items applicable to both the development and evaluation of a prediction model

[2] Separately for all model building approaches.

[3] TRIPOD-Cluster is a checklist of reporting recommendations for studies developing or validating models that explicitly account for clustering or explore heterogeneity in model performance (eg, at different hospitals or centres). Debray et al, BMJ 2023; 380: e071018 [DOI: 10.1136/bmj-2022-071018]

| | | | | |
|---|---|---|---|---|
| *Training versus evaluation* | 16 | D;E | Identify any differences between the development and evaluation data in healthcare setting, eligibility criteria, outcome, and predictors | 8, Table S1 |
| *Ethical approval* | 17 | D;E | Name the institutional research board or ethics committee that approved the study and describe the participant-informed consent or the ethics committee waiver of informed consent | 6 |
| **OPEN SCIENCE** | | | | |
| *Funding* | 18a | D;E | Give the source of funding and the role of the funders for the present study | 2 |
| *Conflicts of interest* | 18b | D;E | Declare any conflicts of interest and financial disclosures for all authors | 2 |
| *Protocol* | 18c | D;E | Indicate where the study protocol can be accessed or state that a protocol was not prepared | 2, 6-7 |
| *Registration* | 18d | D;E | Provide registration information for the study, including register name and registration number, or state that the study was not registered | N/A |
| *Data sharing* | 18e | D;E | Provide details of the availability of the study data | 2 |
| *Code sharing* | 18f | D;E | Provide details of the availability of the analytical code[4] | 2 |
| **PATIENT & PUBLIC INVOLVEMENT** | | | | |
| *Patient & Public Involvement* | 19 | D;E | Provide details of any patient and public involvement during the design, conduct, reporting, interpretation, or dissemination of the study or state no involvement. | 2 |
| **RESULTS** | | | | |
| *Participants* | 20a | D;E | Describe the flow of participants through the study, including the number of participants with and without the outcome and, if applicable, a summary of the follow-up time. A diagram may be helpful. | Figure S1, Figure 1 |
| | 20b | D;E | Report the characteristics overall and, where applicable, for each data source or setting, including the key dates, key predictors (including demographics), treatments received, sample size, number of outcome events, follow-up time, and amount of missing data. A table may be helpful. Report any differences across key demographic groups. | 9-10, Table 1 |
| | 20c | E | For model evaluation, show a comparison with the development data of the distribution of important predictors (demographics, predictors, and outcome). | 10, Figure 2 |
| *Model development* | 21 | D;E | Specify the number of participants and outcome events in each analysis (e.g., for model development, hyperparameter tuning, model evaluation) | 9-10 |
| *Model specification* | 22 | D | Provide details of the full prediction model (e.g., formula, code, object, application programming interface) to allow predictions in new individuals and to enable third-party evaluation and implementation, including any restrictions to access or re-use (e.g., freely available, proprietary)[5] | 8-9, Supplemental Methods |
| *Model performance* | 23a | D;E | Report model performance estimates with confidence intervals, including for any key subgroups (e.g., sociodemographic). Consider plots to aid presentation. | 10-11 Figure 2-3 |
| | 23b | D;E | If examined, report results of any heterogeneity in model performance across clusters. See TRIPOD Cluster for additional details[3]. | 10-11 Figure 3 |
| *Model updating* | 24 | E | Report the results from any model updating, including the updated model and subsequent performance | N/A |
| **DISCUSSION** | | | | |
| *Interpretation* | 25 | D;E | Give an overall interpretation of the main results, including issues of fairness in the context of the objectives and previous studies | 11-12 |
| *Limitations* | 26 | D;E | Discuss any limitations of the study (such as a non-representative sample, sample size, overfitting, missing data) and their effects on any biases, statistical uncertainty, and generalizability | 13-14 |
| *Usability of the model in the context of current care* | 27a | D | Describe how poor quality or unavailable input data (e.g., predictor values) should be assessed and handled when implementing the prediction model | 11-14 |
| | 27b | D | Specify whether users will be required to interact in the handling of the input data or use of the model, and what level of expertise is required of users | 13-14 |
| | 27c | D;E | Discuss any next steps for future research, with a specific view to applicability and generalizability of the model | 13-14 |

From: Collins GS, Moons KGM, Dhiman P, et al. *BMJ* 2024;385:e078378. doi:10.1136/bmj-2023-078378

[4] This relates to the analysis code, for example, any data cleaning, feature engineering, model building, evaluation.
[5] This relates to the code to implement the model to get estimates of risk for a new individual.
